# Estimating differing causal roles of glutamate and GABA genes on brain and behavior in autism

**DOI:** 10.1101/2025.04.11.25325636

**Authors:** Viola Hollestein, Tom Claassen, Jilly Naaijen, Geert Poelmans, I. Hyun Ruisch, Ward De Witte, Christian F Beckmann, Christine Ecker, Sarah Baumeister, Pascal-M Aggensteiner, Tobias Banaschewski, Thomas Bourgeron, Eva Loth, Declan GM Murphy, Julian Tillmann, Tony Charman, Emily J.H. Jones, Luke Mason, Bob Oranje, Rosemary Holt, Sven Bölte, Daniel Brandeis, Anna Kaiser, Steven C. R. Williams, David J. Lythgoe, Jan Buitelaar, Nicolaas A Puts

**Affiliations:** Department of Cognitive Neuroscience, Donders Institute for Brain, Cognition and Behaviour, Radboud University Medical Center, Nijmegen, the Netherlands; Institute for Computing and Information Sciences, Faculty of Science, Radboud University, Nijmegen, The Netherlands; Department of Psychology, Institute of Psychiatry, Psychology & Neuroscience, King’s College, London, London, UK; Department of Child and Adolescent Psychiatry, Psychosomatics and Psychotherapy, University Hospital Frankfurt am Main, Goethe University, Frankfurt, Germany; Department of Child and Adolescent Psychiatry and Psychotherapy, Central Institute of Mental Health, Medical Faculty Mannheim, University of Heidelberg, Mannheim, Germany; German Center for Mental Health, Mannheim, Heidelberg, and Ulm, Germany; Institut Pasteur, Human Genetics and Cognitive Functions Unit, Paris, France; Institute for Translational Neurodevelopment, Institute of Psychiatry, Psychology & Neuroscience, King’s College London, London, UK; Department of Forensic and Neurodevelopmental Sciences, Institute of Psychiatry, Psychology & Neuroscience, King’s College London, London, UK; Medical Research Council (MRC) Centre for Neurodevelopmental Disorders, King’s College, London, London, UK; Roche Pharma Research and Early Development, Neuroscience and Rare Diseases, Roche Innovation Center Basel, F. Hoffmann–La Roche Ltd., Basel, Switzerland; Department of Psychology, Institute of Psychiatry, Psychology, & Neuroscience, King’s College, London, London, UK; Centre for Brain and Cognitive Development, Birkbeck, University of London, Henry Wellcome Building, London, UK; University Medical Center Utrecht, Utrecht University, Utrecht, The Netherlands; Center for Neuropsychiatric Schizophrenia Research (CNSR), Copenhagen University Hospital - Mental Health Services CPH, Glostrup, Denmark; Autism Research Centre, Department of Psychiatry, University of Cambridge, Cambridge, UK; Center of Neurodevelopmental Disorders (KIND), Centre for Psychiatry Research; Department of Women’s and Children’s Health, Karolinska Institutet & Stockholm Health Care Services, Region Stockholm, Stockholm, Sweden; Child and Adolescent Psychiatry, Stockholm Health Care Services, Region Stockholm, Stockholm, Sweden; Curtin Autism Research Group, Curtin School of Allied Health, Curtin University, Perth, Western Australia; Department of Child and Adolescent Psychiatry and Psychotherapy, Central Institute of Mental Health, Medical Faculty, Mannheim/Heidelberg University, Mannheim, Germany; Department of Child and Adolescent Psychiatry and Psychotherapy, Psychiatric Hospital, University of Zurich, Zurich, Switzerland; Center for Integrative Human Physiology, University of Zurich, Zurich, Switzerland; Neuroscience Center Zurich, University of Zurich, Zurich, Switzerland; ETH Zurich, Zurich, Switzerland; Department of Neuroimaging, King’s College London, Institute of Psychiatry, Psychology and Neuroscience, London, United Kingdom; Karakter Child and Adolescent Psychiatry University Center, Nijmegen, the Netherlands

**Author notes:** Corresponding author: Viola Hollestein, Department of Cognitive Neuroscience, Donders Institute for Brain, Cognition and Behavior, Radboud University Medical Center, P.O Box 9101, 6500 HB Nijmegen, The Netherlands.

## Abstract

The excitatory/inhibitory (E/I) imbalance theory suggests that excitatory and inhibitory alterations underlies autism characteristics. However, genetic underpinnings of this imbalance and its impact on brain function and behavior remains unclear. We explored causal links between glutamate and GABA gene-set polygenic scores (PGS) for autism and core autism characteristics, putting particular attention on the restricted-and repetitive behaviors domain by including functional activity (fMRI) during inhibitory control (in the anterior cingulate cortex (ACC) and striatum). Causal links between genes, brain and behavior was evaluated using Bayesian Constraint-based Causal Discovery (BCCD) algorithms, to build causal models of these relationships in a discovery sample (LEAP cohort: autistic = 343, neurotypical = 253) and two generalization cohorts with partially overlapping measures (TACTICS cohort: autistic = 60, neurotypical = 100, Simon Simplex Collection: autistic = 2756). In the discovery sample, we found a causal link between glutamate PGS and core clinical characteristics of autism, particularly the communication domain (Autism Diagnostic Interview-Revised) in autistic participants, with 95% reliability. We did not find links between functional activity during inhibitory control and other measures. For one generalization cohort, we further report on the impact of ^1^H-MRS measures of glutamate, identifying a causal link between GABA autism PGS on ACC glutamate concentrations. Not all links were identified in the generalization cohorts, which may be due to clinical and genetic differences between the cohorts. While our results reinforce the previously found association between glutamate genes and core clinical autism behaviors, task-based functional activity may not be causally related to RRBs.

## Introduction

Autism spectrum disorder (autism) is a heterogeneous neurodevelopmental condition characterized by difficulties in social interaction and communication, restricted repetitive behaviors and altered sensory processing (1). Autism is heritable and affected both by rare genetic variants and common genes, but its etiology is not well understood (2,3). An influential hypotheses of its underlying mechanisms is the excitatory/inhibitory (E/I) imbalance hypothesis, which suggests that chemical alterations of excitatory (predominantly glutamate) and inhibitory (predominantly GABA (γ-aminobutyric acid)) neurotransmission underly autism (4). However, we do not know *how* such alterations may give rise to autism characteristics, and studies investigating glutamatergic and GABAergic functions in the brain have had inconsistent results. This is likely due to several factors, including the different aspects of E/I mechanisms studied across animal models, post mortem and in vivo approaches, differences across study populations, and that various alterations in the brain may lead to similar clinical characteristics (5,6).

Most studies discussing the E/I imbalance in autism have focused on either excitatory or inhibitory measures, rather than investigating both simultaneously, which ignores the complex interactions between them that potentially play a part in developing autistic characteristics. E/I imbalance(s) can arise in various ways, which in turn, may underlie different expressions of autism characteristics (7). Excitation and inhibition are fundamental aspects of brain functioning, and E/I mechanisms exist and interact on a cellular level within individual neurons, between neurons within brain regions, and across the whole brain in communication networks. Genes - or rather, the proteins that they encode - are involved in functions across these levels, and genetic associations between glutamate and GABA communication pathways and behavioral autism characteristics have previously been found, mainly in animal studies but also in humans (8–11). We aimed to assess potential causal associations across several domains by combining genetic approximations of glutamate and GABA, core clinical characteristics of autism, and with particular focus on the restricted and repetitive behavior domain by also including regional functional activity during an inhibitory control task.

Restricted and repetitive behaviors are one of the defining traits of autism. A central executive correlate is inhibitory control difficulty, where increased inhibitory control difficulties contribute to increased repetitive behaviors in autistic individuals (12–14). Inhibitory control can be captured in cognitive tasks such as flanker and stop-signal tasks. Fronto-striatal circuits are known to be involved in regulating inhibitory control, and in vivo measures of alterations in glutamate concentrations in the anterior cingulate cortex (ACC) and striatum have been associated with differences in inhibitory control performance (15–17). Yet, studies investigating inhibitory control in autism have had inconsistent results, where some found differences in performance, or functional brain activation, between autistic and neurotypical participants (12–14) while others have not (18–20). Other studies found differences in functional activity during inhibitory control despite an absence of behavioral differences (19,21,22). These inconsistencies could be due to several factors, for example heterogeneity across autistic individuals, differences across study populations, or varying effects across individuals of E/I imbalance on inhibitory control performance and functional brain activity. We need a better understanding of how E/I imbalance underlies behavioral characteristics of autism and affect its underlying brain activity, to disentangle the etiologies of different autism characteristics.

All in all, links across genetic contributions, functional activity and behavioral characteristics in autism are not well understood, and findings so far have had inconsistent approaches, study populations, and results. We aimed to address this by building causal discovery models to evaluate links between measures to identify the most likely causal relationships. This is a data-driven approach that estimates the most likely causal structure between the different measures, which has the potential to direct future investigations more effectively. We also included the possibility that other commonly co-occurring conditions, such as ADHD and anxiety, as well as age and sex may affect these relationships.

More specifically, we used Bayesian Constraint-based Causal Discovery (BCCD), a state-of-the-art algorithm that learns causes and effects from observational data, and detects whether the dependency between variables is direct or mediated through other variables (23). Genetic variation within glutamate and/or GABA pathways was estimated using gene-set polygenic scores, to aggregate the various contributions of these genes. These scores were evaluated for causal relationships with core behavioral characteristics of autism, and brain activity in selected regions of interest during inhibitory control. We used a large sample (N = 596, autistic = 343, neurotypical = 253) as our discovery sample, and two generalization samples (first sample: N = 160, autistic = 60, neurotypical = 100, second sample: N autistic = 2756). These additional cohorts did not provide identical measures and age ranges as the discovery sample, and were therefore useful for generalization analyses. The first generalization sample additionally included ^1^H-MRS measures of in vivo glutamate concentrations in ACC and striatum, which allowed for inclusion of another level of E/I proxies to be evaluated with the genetic and functional magnetic resonance imaging (fMRI) based measures.

BCCD differs from commonly used regression analyses as it disentangles causal structures, while regression analyses test strengths of presupposed associations under the assumption that such relationships are true. By identifying the most plausible structures between data modalities, we could identify which relationships are most likely useful to focus on in future testing.

## Materials and Methods

### Participants

Data from three cohorts were used, one as a discovery sample and two others as replication samples. Our discovery sample was the Longitudinal European Autism Project (LEAP) cohort, part of the AIMS-2-TRIALS research programme (https://www.aims-2-trials.eu/) (24–26). We used data from 596 participants (autistic = 343, neurotypical = 253) aged 6 to 30 years, collected at six centers across Europe (Institute of Psychiatry, Psychology and Neuroscience, King’s College London (IoPPN/KCL, UK), Autism Research Centre, University of Cambridge (UCAM, UK), University Medical Centre Utrecht (UMCU, Netherlands), Radboud University Medical Centre (RUMC, Netherlands), Central Institute of Mental Health (CIMH, Germany), and the University Campus Bio-Medico (UCBM) in Rome, Italy).

The first generalization sample was from the European Union funded TACTICS cohort (www.tactics-project.eu) (27), where we included data from of 160 participants (autistic = 60, neurotypical = 100), aged 8 to 13 years old, collected from three centers across Europe (Radboud University Medical Centre, Nijmegen, The Netherlands; King’s College London, London, United Kingdom; and Central Institute of Mental Health, Mannheim, Germany). Details regarding inclusion and exclusion criteria for both cohorts can be found in the Supplementary material.

The second generalization sample included genetic and behavioral measures from the Simons Simplex Collection (SSC), where we used data from 2756 autistic participants between 4 and 18 years old, collected in the USA (28).

### Phenotypic measures

The phenotypic measures in the LEAP cohort were part of a larger test battery (see (24)). We included three questionnaires capturing the core autism characteristics; social behaviors (Social Responsiveness Scale-Revised (SRS-2; (29)), repetitive behaviors (Repetitive Behavior Scale-Revised (RBS-R; (30) and sensory processing (Short Sensory Profile (SSP; (31)). In the autistic participants, the autism scores on the Autism Diagnostic Observation Schedule Second Edition (ADOS-2, (32)) and Autism Diagnostic Interview - Revised (ADI-R, (33)) were available. The RBS-R and ADI-R were also available in the TACTICS cohort. The Children’s Social Behavior Questionnaire (CSBQ; (34)) in the TACTICS sample, similar to the SRS-2 used in LEAP, was included as a similar measure of social communicative behaviors. These questionnaires were either parent-or self-report depending on age and diagnostic group. In the SSC cohort, the SRS-2, RBS-R, ADOS-2 and ADI-R were available. An overview of what measures were used in which cohort can be found in Supplementary Table S1.

We included measures of the most common co-occurring conditions to account for potential confounding or mediating effects between our measures of interest; ADHD (DSM-5 ADHD-Rating Scale), anxiety (Beck Anxiety Inventory (BAI; (35)), and depression (Beck Depression Inventory-II; (36)) in the LEAP cohort. In the TACTICS cohort, ADHD measures from a different rating scale were also available (Conners’ Parent Rating Scale (CPRS-R; (37)).

### Genetics

#### Genotyping

Genotyping of the LEAP cohort was performed at the Centre National de Recherche en Génomique Humaine (CNRGH) using the Infinium OmniExpress-24v1 BeadChip Illumina. Genotyping of the TACTICS cohort was performed using the PsychChip_v1-1_15073391 platform in Bonn. For details on how these were performed, see the Supplementary material.

#### Gene-set selection

The glutamate (n = 72) and GABA (n = 124) gene-sets have been used in several studies previously (9,38,39), and consist of genes encoding proteins involved in glutamatergic and GABAergic pathways in the brain. The gene selection was based on Ingenuity Pathway Analysis software (http://www.ingenuity.com), a database for genetic pathway analysis based on evidence from scientific literature and other sources such as gene expression and annotation databases, assigning genes to groups and categories of functionally related genes. The complete lists of genes in each gene-set can be found in Supplementary Tables S2-S3.

#### Gene-set polygenic scores

We derived gene-set based polygenic scores (PGS) for our glutamate and GABA gene-sets using the PRSet function in PRSice-2 (40,41) with the summary statistics of the PGC ASD GWAS (Genome-Wide Association Study) (42). SNPs were clumped based on LD, using PRSice default settings (bidirectional 250Kb-window and R2-threshold of 0.1), resulting in 103.045 LD-clumped SNPs in the LEAP cohort, 103.043 LD-clumped SNPs in the TACTICS cohort, and 174.617 LD-clumped SNPs in the SSC cohort. Glutamate and GABA gene-set PGS were calculated at a p-value threshold of 1, to include the whole gene-set in the PGS.

### Neuroimaging

#### fMRI acquisition

In both the LEAP and TACTICS cohorts, all sites acquired data on 3 T Magnetic Resonance (MR) scanners, obtaining functional MRI during an inhibitory control task (see below for details). Additional Proton Magnetic Resonance Spectroscopy (^1^H-MRS) data were acquired in the TACTICS cohort to measure glutamate concentrations in the ACC and striatum. The scanner and sequence details for both cohorts, as well as the processing details for the ^1^H-MRS data, can be seen in the Supplementary material and Tables S4-S5.

#### fMRI inhibitory control tasks

In the LEAP cohort, the inhibitory control task was a modified version of a combined flanker-go/nogo task (43). In the TACTICS cohort, the inhibitory control task was a stop-signal task (44). The data from both cohorts have been preprocessed and analyzed previously, and were reused here for consistency with prior work. Task contrast processing was performed identically in both cohorts using SPM12 (Statistical Parametric Mapping release 12, https://www.fil.ion.ucl.ac.uk/spm/) to allow for comparisons between the two inhibitory control tasks across LEAP and TACTICS. Contrasts reflecting successful and failed inhibitory control were therefore modified from standard contrasts in the LEAP task, detailed in the Supplement. With the created contrasts we extracted the mean beta weights, which were the estimated changes in BOLD activity during our inhibitory control contrasts, from the ACC and striatum. These regions were our task-relevant regions of interest for the inhibitory control task, and additionally have ^1^H-MRS measures of glutamate from ACC and striatum in the TACTICS cohort. Registration between fMRI and ^1^H-MRS was done with the MarsBar toolbox (45), using the voxel placement of the ^1^H-MRS measures as the region of interest (ROI) in both cohorts. This resulted in four estimations of mean beta weight of functional activity for each participant, for each contrast (successful and failed inhibitory control), and in each brain region (ACC and striatum). The LEAP cohort had fMRI data available from 354 participants. The TACTICS cohort had fMRI data available from 44 participants who additionally had ^1^H-MRS measures of glutamate concentrations in ACC and striatum.

## Statistical analysis

### BCCD analysis

We used the Bayesian constraint-based causal discovery (BCCD) algorithm to find direct and indirect (mediated by other variables) interactions (23). The benefits of the BCCD algorithm are its ability to handle a combination of continuous and discrete variables, while also handling missing data, which is dealt with when estimating the correlation matrix using expectation maximization algorithms (46). This method combines the strengths of constraint-based methods giving strong and clear causal relationships, and of score-based methods estimating confidence measures of inferred causal relationships. Under a set of assumptions, the BCCD algorithm looks for combinations of dependence and independence patterns in the data, which may imply that a variable is likely to cause another variable. BCCD gives us reliable estimated causal relationships between variables, with an estimation of the likelihood of these relationships. It has been evaluated and confirmed to be an effective method in this context, and has been used on similar types of data sets, investigating other neurodevelopmental conditions such as ADHD, and psychopathologies (47–51).

This is a hypothesis-free approach, based on a set of assumptions such as no cyclic dependencies being present (for more details, see (23,50)), and can therefore validate previously found associations between data modalities. It also provides additional information over regression-based approaches for causal interpretation. In contrast to regression analysis, we here do not assume a model, but generate a quantified causal model that best explains the observed structure between the data. The observational data fed into the BCCD is mapped through a Gaussian transformation into a correlation matrix (23), followed by an efficient search to obtain Bayesian reliability scores, resulting in weighted independence constraints. Lastly, the logical independence constraints are used with initially defined background knowledge (behavioral measures cannot cause polygenic scores, sex, or age) and creates an output model. Estimated causal links with a reliability of 60% or higher are considered robust, which are presented in Figures 1-5. BCCD does not provide effect sizes, unlike regression models which estimate effects under assumed relationships. We here provide likelihood estimations of the identified relationships, which can be found across all models in Supplementary Tables S9-S18.

**Figure 1.**
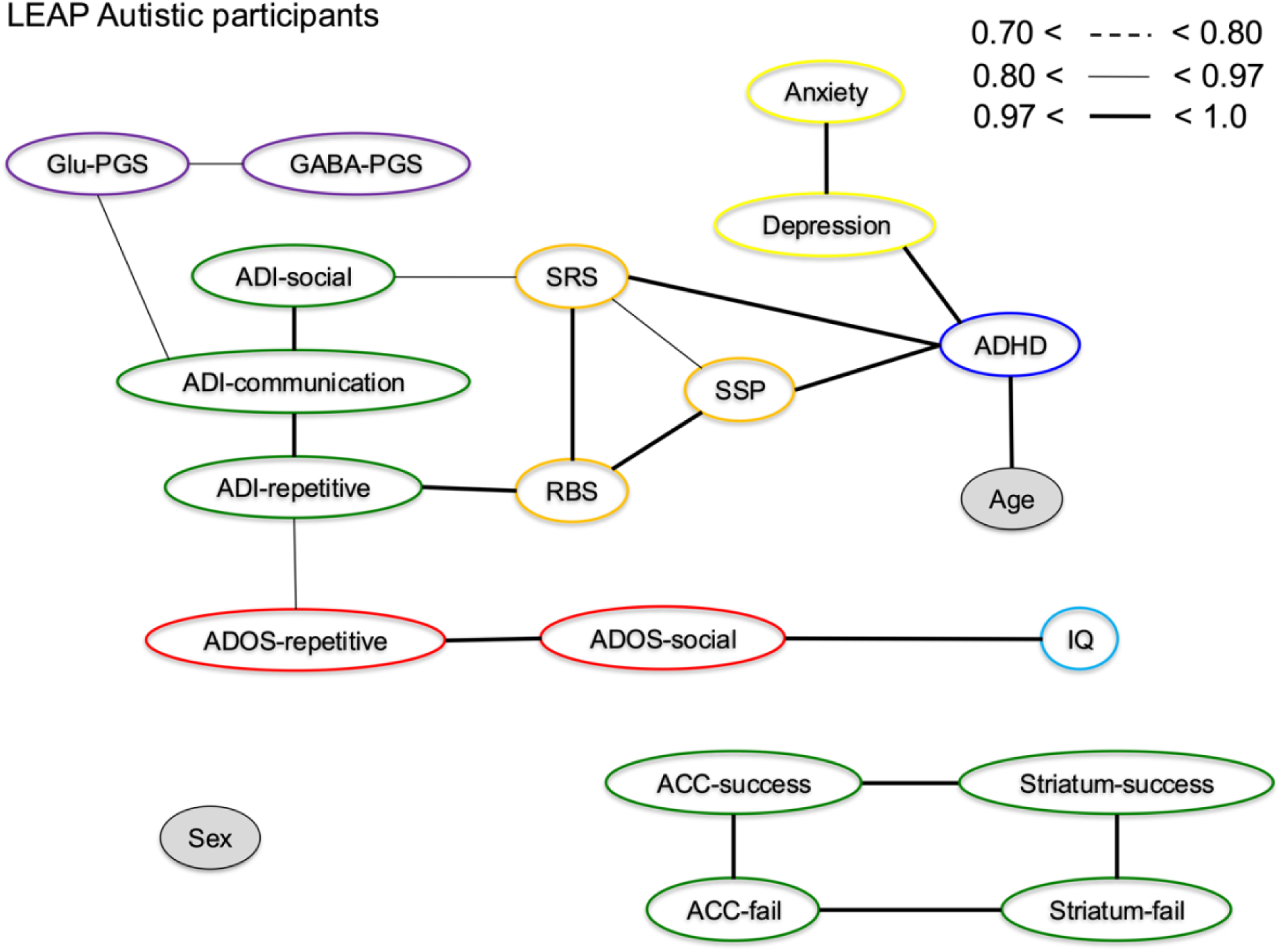
BCCD LEAP Autistic participants. Output causal model representing causal relationships between the genetic, task-based functional MRI and behavioral measures. Reliability estimates for edges shown here are depicted as ranges of percentages as defined in the figure. Glu-PGS, Glutamate polygenic score for autism; GABA-PGS, GABA polygenic score for autism; SRS-2, Social Responsiveness Scale-Revised, RBS-R, Repetitive Behavior Scale-Revised; SSP, Short Sensory Profile; ADI-social, Autism Diagnostic Interview-Revised Social domain; ADI-communication, Autism Diagnostic Interview-Revised Communication domain; ADI-repetitive, Autism Diagnostic Interview-Revised Restricted and Repetitive Behaviors domain; ADOS social, Autism Diagnostic Observation Schedule Second Edition Social affect; ADOS repetitive, Autism Diagnostic Observation Schedule Second Edition Restricted and Repetitive Behaviors; ADOS-total, Autism Diagnostic Observation Schedule Second Edition Total score; Anxiety, Beck Anxiety Inventory; Depression, Beck Depression Inventory; ADHD, DSM-V ADHD Rating Scale; ACC-success, BOLD signal in ACC during successful inhibitory control; Striatum-success, BOLD signal in striatum during successful inhibitory control; ACC-fail, BOLD signal in ACC during failed inhibitory control; Striatum-fail, BOLD signal in striatum during failed inhibitory control.

To investigate potential differences between autistic and neurotypical participants, we created separate models with autistic and neurotypical LEAP participants, while additionally creating a model with all participants combined. As the SSC cohort only consists of autistic participants, separating by diagnostic group allowed for a more direct comparison between the cohorts.

For each model (autism, neurotypical, and whole cohort), participants with > 50% of the data missing were excluded to reduce the risk of unwanted imputation effects, resulting in 596 LEAP participants (autistic = 343, neurotypical = 253) and 2756 SSC participants. This threshold was set to balance minimizing possible imputation impact, and maximizing available information from collected data. In the TACTICS cohort, a large number of participants had just over > 50% missing data, and we therefore included participants with up to 60% missing data to adjust this balance, resulting in 160 included participants (autistic = 60, neurotypical = 100). This did not affect the estimated causal structure in the model, but provided increased power for more accurate model estimation. For an overview of which measures were included in what cohort, see Supplementary Table S1.

### Comparing cohorts

Post-hoc tests were performed to compare gene-set PGS and ADI-R scores between the LEAP and SSC cohorts using standard two-sided t-tests in base R software (52).

## Results

### Demographics

Demographic and clinical characteristics of all cohorts are shown in Tables S6-S8 in the Supplement. In the LEAP cohort, no differences were found between the diagnostic groups in age or sex, the autism group had a lower IQ compared to neurotypical participants (details can be seen in Table S6). In the TACTICS cohort, there were no group differences in age, female-to-male ratio, or IQ. The SSC cohort only included autistic participants, therefore no group comparisons were performed.

### Bayesian constraint-based causal discovery

The models output by the BCCD algorithm can be seen in the figures below, where variables of interest (node) are connected via lines (edges), representing an estimated causal relationship. The figures show edges with a causal link reliability of >= 60%. Exact values of all edges and estimated correlations between variables can be seen in Supplementary Tables S9-S18.

*LEAP:* Starting with LEAP, Figure 1 shows the autism group, Figure 2 the neurotypical group and Figure 3 the whole cohort. Only Figure 1 includes the measures of ADOS-2 and ADI-R, as these were only measured in the autistic participants.

**Figure 2.**
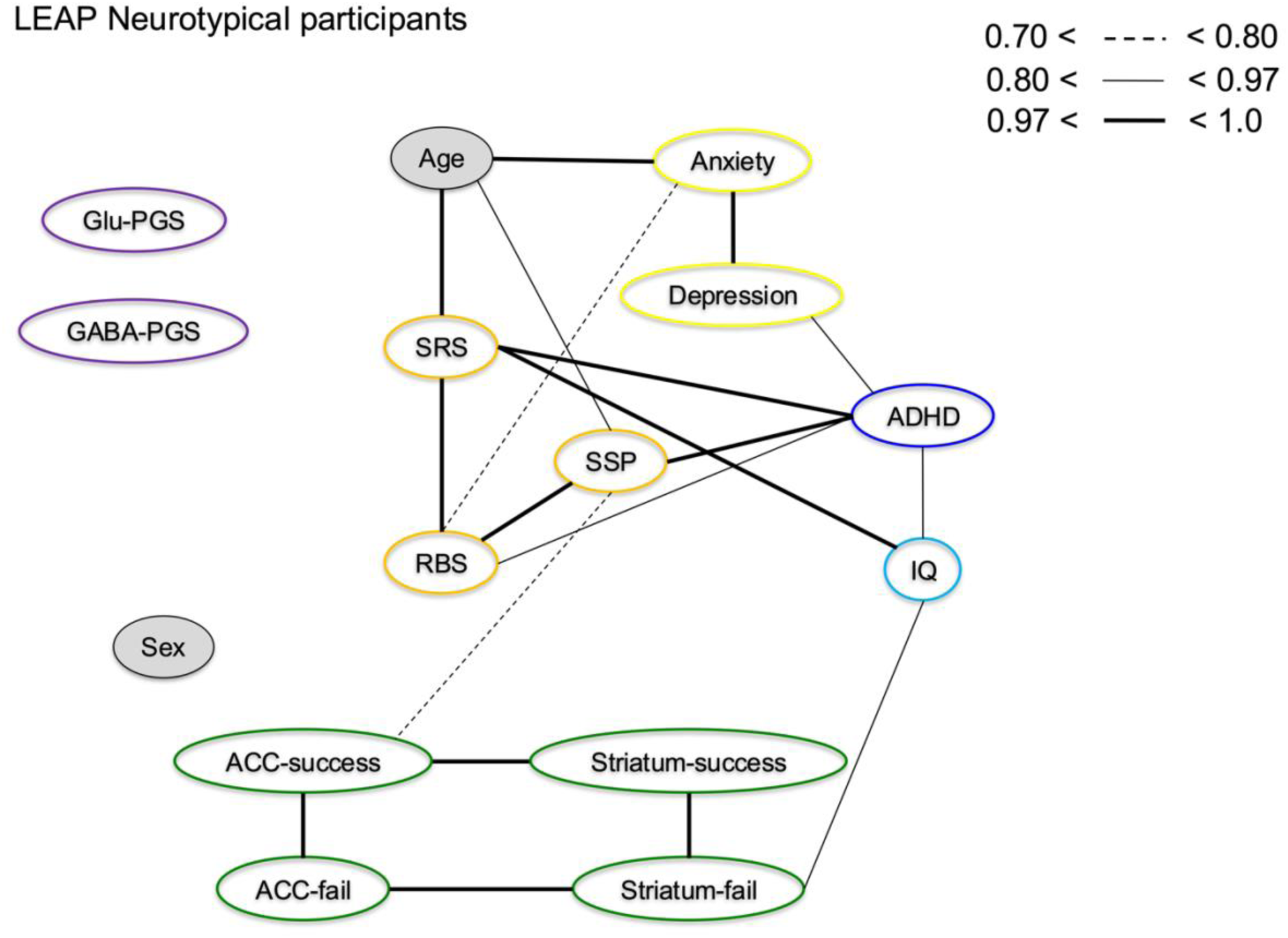
BCCD LEAP Neurotypical participants. Output causal model representing causal relationships between the genetic, task-based functional MRI and behavioral measures. Reliability estimates for edges shown here are depicted as ranges of percentages as defined in the figure. Glu-PGS, Glutamate polygenic score for autism; GABA-PGS, GABA polygenic score for autism; SRS-2, Social Responsiveness Scale-Revised, RBS-R, Repetitive Behavior Scale-Revised; SSP, Short Sensory Profile; Anxiety, Beck Anxiety Inventory; Depression, Beck Depression Inventory; ADHD, DSM-V ADHD Rating Scale; ACC-success, BOLD signal in ACC during successful inhibitory control; Striatum-success, BOLD signal in striatum during successful inhibitory control; ACC-fail, BOLD signal in ACC during failed inhibitory control; Striatum-fail, BOLD signal in striatum during failed inhibitory control.

**Figure 3.**
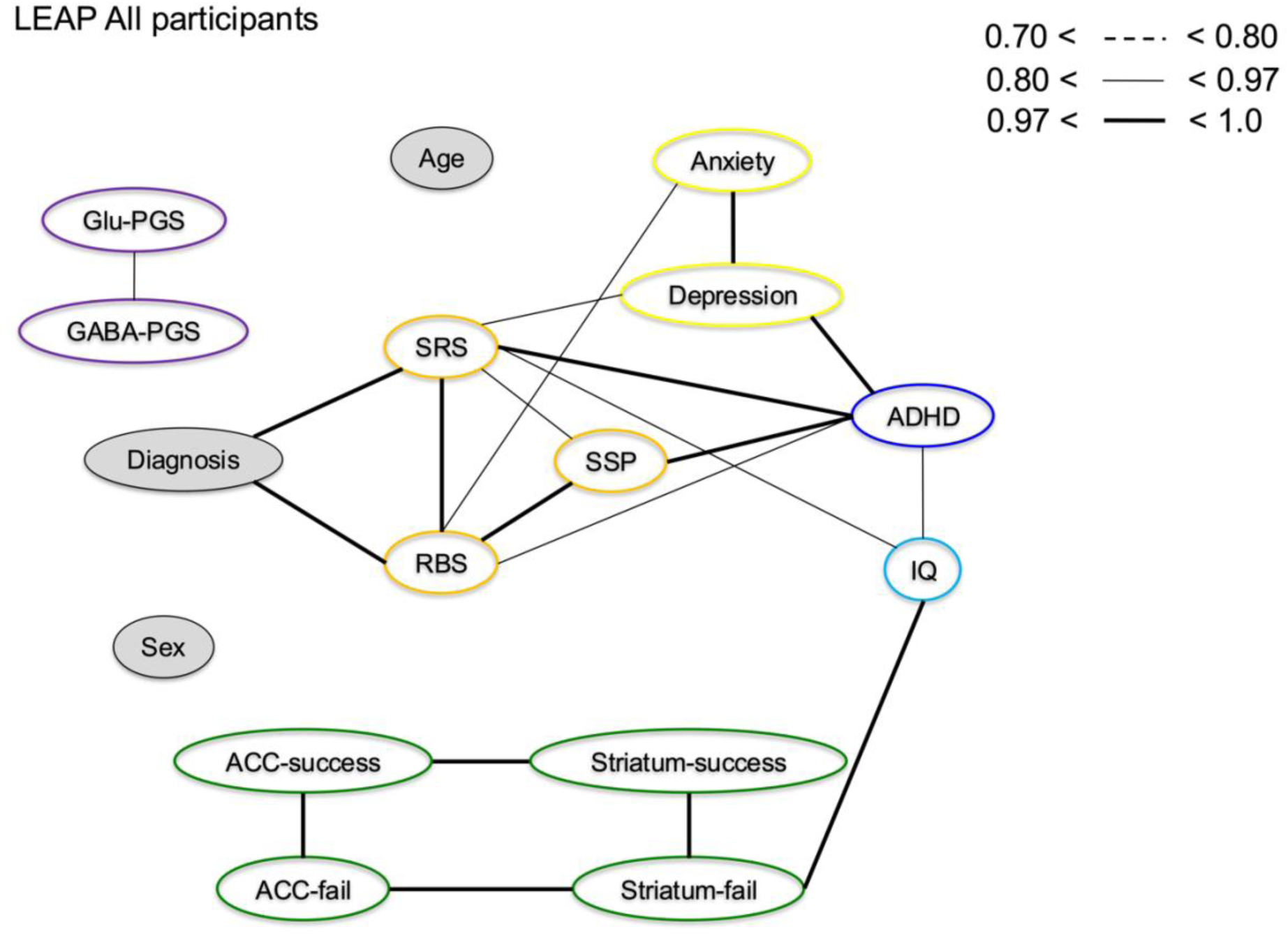
BCCD LEAP All participants. Output causal model representing causal relationships between the genetic, task-based functional MRI and behavioral measures. Reliability estimates for edges shown here are depicted as ranges of percentages as defined in the figure. Glu-PGS, Glutamate polygenic score for autism; GABA-PGS, GABA polygenic score for autism; SRS-2, Social Responsiveness Scale-Revised, RBS-R, Repetitive Behavior Scale-Revised; SSP, Short Sensory Profile; ADI-social, Autism Diagnostic Interview-Revised Social domain; ADI- communication, Autism Diagnostic Interview-Revised Communication domain; ADI-repetitive, Autism Diagnostic Interview-Revised Restricted and Repetitive Behaviors domain; ADHD, DSM-V ADHD Rating Scale; ACC-success, BOLD signal in ACC during successful inhibitory control; Striatum-success, BOLD signal in striatum during successful inhibitory control; ACC- fail, BOLD signal in ACC during failed inhibitory control; Striatum-fail, BOLD signal in striatum during failed inhibitory control.

In the autism group (Figure 1), we observed a direct causal link of 95% reliability (Supplementary Table S9) between the glutamate PGS and the ADI-R communication domain. Additionally, the ADI-R communication domain was strongly linked to the social and repetitive domains as well (at least 97% reliability, Table S9), indicating that a glutamate PGS to ADI-R link is primarily mediated by the communication domain. The glutamate and GABA PGS were causally linked to each other with 97% reliability in the autism group, and this link was not present in the neurotypical group (Figure 2). As the glutamate and GABA PGS are both genetic scores, we could not infer directionality between them.

Across the whole cohort (Figure 3), as well as in the separate groups (Figures 1 and 2), we observed causal links between RBS-R, SRS-2 and SSP scores. These results show that what are typically referred to as the core clinical behaviors of autism (repetitive behaviors, social-communicative behaviors and sensory processing) are not just related within autistic individuals but that these behaviors affect each other across participants regardless of diagnosis. In the autism group (Figure 1) we also observed links between SRS-2 and the ADI-R social domain, and between RBS-R and the ADI-R repetitive behavior domain, confirming that the SRS-2 and RBS-R questionnaires capture similar behavioral traits as the ADI-R social and repetitive domains.

The BOLD contrast measures of functional activity during successful and failed inhibitory control in the ACC and striatum were causally connected with at least 97% reliability, but were not associated with other measures. This is seen as they are separate from the other measures in the models, both across the whole cohort and in the separate groups (Figures 1-3, Supplementary Tables S9, S11, S13). In the neurotypical group however, there was a causal link of 93% reliability (Supplementary Table S11) between IQ and failed inhibitory control BOLD activity in striatum, which was not present in the autism group. This link was also seen across the whole sample (Figure 3).

*TACTICS:* We did not have enough data to divide into diagnostic groups, and therefore only used the whole sample (Figure 4). We did however replicate some of the causal relationships between the behavioral measures that overlap in the LEAP cohort. Firstly, the CSBQ, an equivalent measure to the SRS in LEAP, showed causal links to RBS and ADHD scores similar to the LEAP cohort (Figure 3), indicating that the causal links between social and repetitive behaviors captured by these questionnaires are robust. Secondly, we replicated the link between BOLD signal in striatum during inhibitory control and IQ as seen in the LEAP cohort, particularly in the neurotypical group. The BOLD activity during failed and successful inhibitory control was not causally linked in the same way as in the LEAP cohort, however, there was a mediating effect by age between successful inhibitory control in ACC and striatum which potentially points towards the differences in age ranges across the TACTICS and LEAP cohorts.

**Figure 4.**
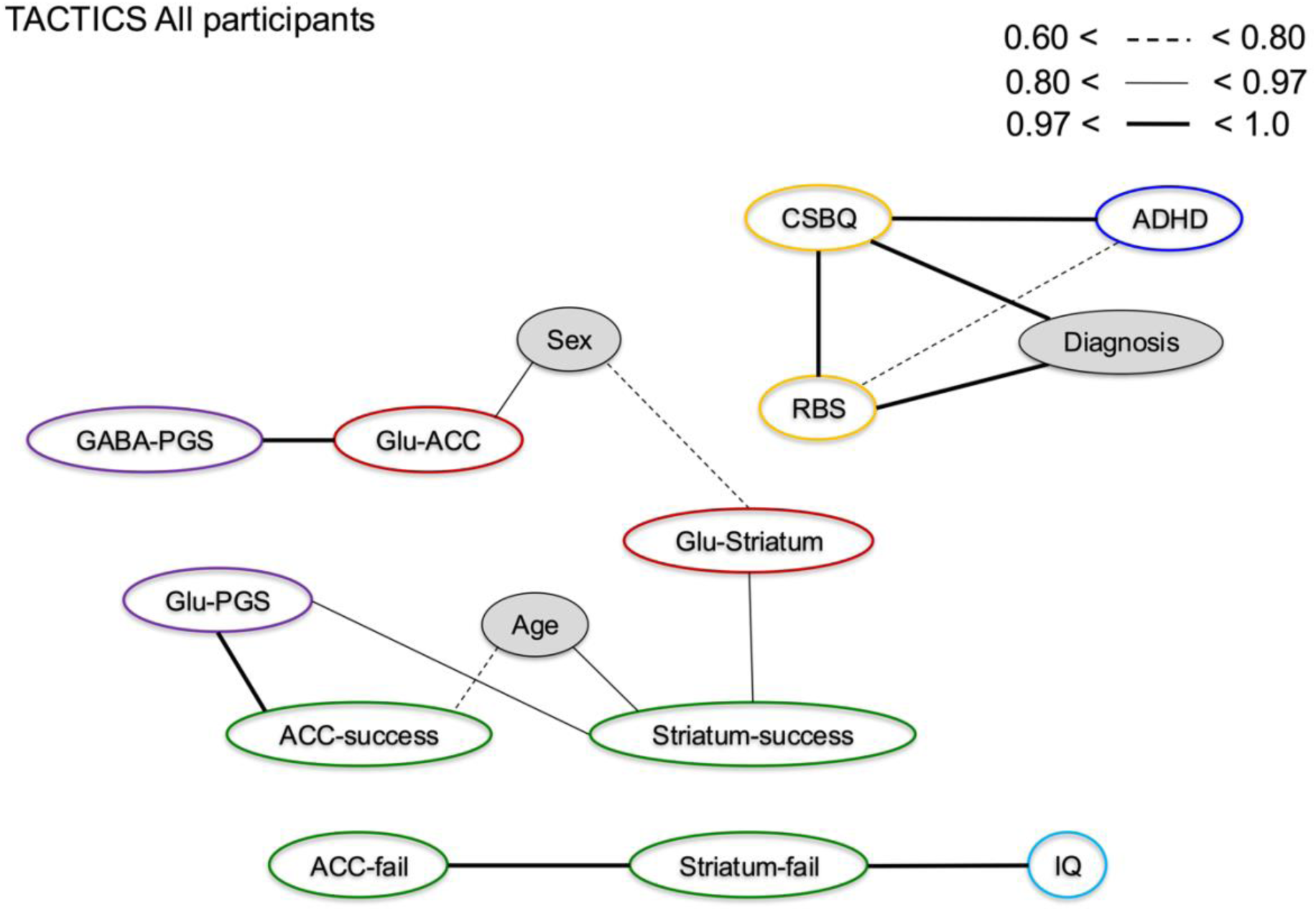
BCCD TACTICS All participants. Output causal model representing causal relationships between the genetic, task-based functional MRI and behavioral measures. Reliability estimates for edges shown here are depicted as ranges of percentages as defined in the figure.

**Figure 5.**
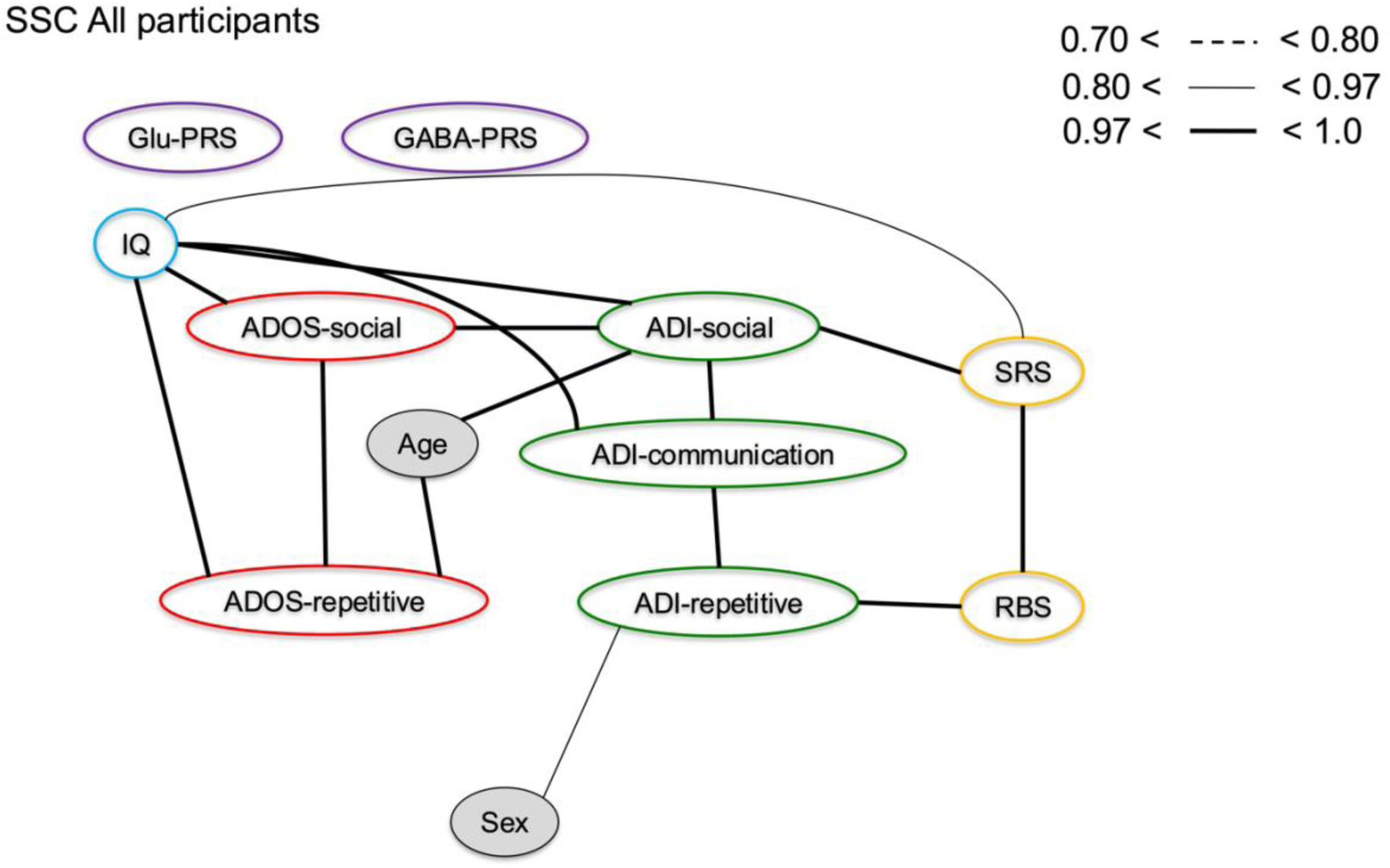
BCCD SSC Autistic participants. Output causal model representing causal relationships between the genetic and behavioral measures. Reliability estimates for edges shown here are depicted as ranges of percentages as defined in the figure. Glu-PGS, Glutamate polygenic score for autism; GABA-PGS, GABA polygenic score for autism; SRS-2, Social Responsiveness Scale-Revised, RBS-R, Repetitive Behavior Scale-Revised; ADI-social, Autism Diagnostic Interview-Revised Social domain; ADI-communication, Autism Diagnostic Interview-Revised Communication domain; ADI-repetitive, Autism Diagnostic Interview-Revised Restricted and Repetitive Behaviors domain; ADOS social, Autism Diagnostic Observation Schedule Second Edition Social affect; ADOS repetitive, Autism Diagnostic Observation Schedule Second Edition Restricted and Repetitive Behaviors; ADOS-total, Autism Diagnostic Observation Schedule Second Edition Total score.

Looking at the addition of ^1^H-MRS glutamate concentrations in this model, striatal glutamate concentrations had a causal link to striatal BOLD activity during successful inhibitory control with 96% reliability (Supplementary Table S15). GABA PGS showed a causal link with 99% reliability to ACC glutamate (Supplementary Table S15). To exclude that these results were introduced due to imputation effects, as we included participants with up to 60% missing data in this cohort, we confirmed that these patterns were also present when running the model without imputation. We did not identify causal links between the glutamate and GABA PGS with behavioral measures of autism characteristics in this cohort.

As the sample size in TACTICS was relatively small we wanted to attempt to replicate, or generalize, our findings in another cohort, being especially interested in the gene-set PGS links to behavioral measures in the autism group in the LEAP sample. For this, we used the SSC, the demographic information of the SSC is available in the Supplementary Table S8.

*SSC:* Figure 5 shows causal links between the SRS-2 and ADI-R social domain, and with the RBS-R and ADI-R repetitive domain, replicating the behavioral links between these in the LEAP cohort. However, the reported causal link between glutamate PGS to ADI-R in LEAP were not captured here, and glutamate and GABA PGS were also not causally linked to each other, while they were in the autism group in the LEAP cohort.

### Comparing cohorts

To disentangle why some estimated causal links did not generalize across cohorts, we compared behavioral and genetic profiles between the LEAP and SSC cohorts using t-tests between the autistic samples on the glutamate and GABA PGS and ADI-R domains. These tests showed that the cohorts differ from each other in their glutamate and GABA PGS, and the ADI-R measures (all p-values <0.001), indicating a difference in the genetic and clinical profiles of the SSC cohort compared to our European LEAP cohort. These results can be seen in the Supplementary Table S19, and individual data points can be seen in Supplementary Figures S2-S5.

Glu-PGS, Glutamate polygenic score for autism; GABA-PGS, GABA polygenic score for autism; CSBQ, Children’s Social Behavior Questionnaire; RBS-R, Repetitive Behavior Scale-Revised; SSP, Short Sensory Profile; ADI-social, Autism Diagnostic Interview-Revised Social domain; ADI-communication, Autism Diagnostic Interview-Revised Communication domain; ADI-repetitive, Autism Diagnostic Interview-Revised Restricted and Repetitive Behaviors domain; ADHD, Conners’ Parent Rating Scale; ACC-success, BOLD signal in ACC during successful inhibitory control; Striatum-success, BOLD signal in striatum during successful inhibitory control; ACC-fail, BOLD signal in ACC during failed inhibitory control; Striatum-fail, BOLD signal in striatum during failed inhibitory control.

## Discussion

We used BCCD to identify probable causal relationships between glutamate and GABA polygenic scores (PGS) behavioral measures of autism traits and fMRI, in one discovery cohort and two generalization cohorts with partially overlapping measures. We did not observe links between functional activity during inhibitory control with genetic or behavioral measures, but identified plausible causal relationships between genetic and behavioral measures. We observed a causal connection between glutamate (autism) PGS and ADI-R in the autism group in LEAP, primarily to the communication domain, suggesting shared genetics between autism polygenic scores and autism symptoms captured by the ADI-R. This confirms previous findings in LEAP that used aggregated genetic variation rather than PGS (9). Additionally, we found a causal link between the GABA (autism) PGS and ACC glutamate in TACTICS, supporting earlier work showing reduced ACC glutamate in autistic, compared to neurotypical, participants (15). These causal links show that glutamate and GABA genes causally underlie autism traits in distinct ways, and are informative for future work to disentangle regional specificity and identify more specific biological underpinnings of these causal relationships, and stratify individual differences.

In the autism LEAP sample, glutamate and GABA PGS were causally linked. This may reflect interactions between glutamate and GABA communication pathways affecting autism likelihood in autistic individuals. It may also reflect a shared, unobserved, variable causing this link, or a selection effect of the autism sample in this cohort, which are factors considered by the BCCD algorithm. However, these findings were absent in the SSC data, discussed below. Glutamate and GABA are metabolically closely related and interact as part of neuronal functioning (53,54). Causal interactions between glutamate and GABA in the autism group specifically may therefore reflect compensatory mechanisms of excitatory and inhibitory functions attempting to maintain homeostasis (5). Although the diagnostic groups are not compared directly, this indicates different relationships between the autism polygenic scores in these glutamate and GABA gene-sets between autistic and neurotypical participants.

The estimated causal links between genetic and behavioral measures found here are novel and important for understanding the etiology of autism, but are relatively far removed from the mechanisms in the brain that we try to disentangle, as we do not pick up on e.g. ratios between excitation and inhibition, or regional specificity of where in the brain differences are expressed. Causal structures estimated between these data modalities do not necessarily reflect direct causal relationships, as there are many biological processes between genetics and behavior that were not captured in the measures included. These relationships nevertheless reflect causal effects of glutamate and GABA genes on behavior, and future work should include additional measures such as in vivo ^1^H-MRS measures of glutamate and GABA combined, to investigate how ratios between these excitatory/inhibitory measures may indicate (im)balances and how they relate to other brain, gene and behavior measures.

We started to bridge this gap by including glutamate ^1^H-MRS measures in the TACTICS cohort, although we did not observe links mediating the relationships between genes and behavior, or to brain activity during inhibitory control. There were, however, links between the successful and failed inhibitory control contrasts. The contrasts in the ACC and striatum had strong links across both cohorts, LEAP and TACTICS, although the structure of links between failed and successful inhibitory control were not. This is possibly due to LEAP and TACTICS using different inhibitory control tasks, or the demographic differences between the cohorts. While the contrasts of successful and failed inhibitory control were created to be as identical across the cohorts as possible, the potential differences across the tasks constitute a limitation for attempts to replicate and generalize across the cohorts. Further, the ACC and striatum BOLD signals were relatively separate from the other measures (Figures 3-4) suggesting that the gene-set autism PGS have little effect on functional activity contrasts in these brain regions. It should also be noted that the associations of glutamate genes could indicate both increased and decreased glutamate function, but broadly shows that genetic disposition towards differences in glutamate impacts the development of autistic traits and thus, that differences in glutamate function, are causally driving autism characteristics. This is consistent with prior work showing that altered concentrations of both glutamate and GABA are associated with autism, although these neurotransmitters have rarely been investigated simultaneously (10,15,16,55–62).

Across behavioral measures, we found robust and consistent causal relationships between several behavioral measures that generalize across the LEAP, TACTICS and SSC cohorts. In particular, the SRS-2 had a strong causal link with the ADI-R social score, and the RBS-R with the ADI-R repetitive score in both LEAP and SSC. This confirms that these measures capture similar aspects of social and repetitive autism traits and symptoms, reinforcing associations established previously (24) and validating them using a hypothesis-free, data driven approach. Identifying these relationships across the three cohorts also gave us strong confidence that the models themselves are robust and that other findings could be considered reliable.

The glutamate PGS to ADI-R relationships were not replicated in the SSC cohort. Genetics, including polygenic scores, cannot fully explain complex behaviors as they aggregate the small effects of common genetic variants. Polygenic scores therefore do not capture all potential factors where genetics may affect autism likelihood or expression of specific traits. The glutamate PGS to ADI-R relationships may still exist in the SSC cohort, but be operationalized differently and mediated by factors not included in the model, such as epigenetic or environmental factors. The varying results may also be due to differences in genetic and clinical profiles of autism traits in the SSC compared to LEAP. Our post-hoc tests showed differences in the glutamate and GABA PGS. While the GWAS used to create the PGS is the largest available to date, it is based on a European cohort, which may be less accurate for the USA SSC data (63). Furthermore, the PRSet tool used to calculate the PGS is better powered in larger sample sizes (40). SSC also differs in its clinical profile, as seen in the higher ADI-R scores. This is likely due to more stringent inclusion criteria in the SSC cohort, where ADI-R and ADOS-2 cutoff for diagnosis were inclusion criteria. The LEAP and TACTICS cohorts instead relied on prior clinical diagnosis, and used diagnostic scores for ADOS-2 and ADI-R as an additional validation rather than inclusion criterion, which potentially led to subtle differences in the recruitment of participants. However, we do replicate the causal relationships between the behavioral measures in the SSC cohort. The differences across cohorts are relevant and warrant further investigation. They also highlight the need for caution when generalizing findings beyond datasets like these, particularly outside Europe and the USA, where additional cultural and clinical variations may exist. Such nuances, even between large cohorts like LEAP and SSC can have important impacts on results. It is clear that we cannot solely rely on large sample sizes to combat this.

In conclusion, we found putatively causal relationships between glutamate PGS for autism with behavioral autism traits as captured by the ADI-R, which were not seen with the GABA PGS. In another cohort (TACTICS), we identified a likely causal link between GABA PGS for autism with ACC glutamate concentrations. Glutamate and GABA genes show different roles underlying behavioral autistic characteristics, potentially affecting different assessment levels as captured here with the glutamate metabolite measures. This is informative for future research disentangling more specific biological underpinnings of these relationships and how it underlies autism characterstics.

## Data Availability

LEAP: All data in the present study will be available upon request to the consortium in the future

## Acknowledgments and Funding

LEAP: The results leading to this publication have received funding from the Innovative Medicines Initiative 2 Joint Undertaking under grant agreement No 777394 for the project AIMS-2-TRIALS. This Joint Undertaking receives support from the European Union’s Horizon 2020 research and innovation programme and EFPIA and AUTISM SPEAKS, Autistica, SFARI. Any views expressed are those of the author(s) and not necessarily those of the funders (IHI-JU2).

TACTICS: The research leading to these results was supported by the European Community’s Seventh Framework Program (FP7/2007-2013) TACTICS under grant agreement no. 278948; the Innovative Medicines Initiative Joint Undertaking under grant agreement number 115300 (EU-AIMS), resources of which are composed of financial contribution from the European Union’s Seventh Framework Programme (FP7- /2007 - 2013) and the European Federation of Pharmaceutical Industries and Associations (EFPIA) companies’ in kind contribution.

## Conflict of interest statements

Prof. Banaschewski served in an advisory or consultancy role for ADHS digital, Infectopharm, Lundbeck, Medice, Neurim Pharmaceuticals, Oberberg GmbH, Roche, and Takeda. He received conference support or speaker’s fee by Medice and Takeda. He received royalties from Hogrefe, Kohlhammer, CIP Medien, Oxford University Press; the present work is unrelated to these relationships. Prof. Buitelaar has been in the past 3 years a consultant to / member of advisory board of / and/or speaker for Takeda, Roche, Medice, Angelini, Janssen, Boehringer-Ingelheim, and Servier. He is not an employee of any of these companies, and not a stock shareholder of any of these companies. He has no other financial or material support, including expert testimony, patents, royalties. Dr. Poelmans is director and Dr. Ruisch. and Dr. de Witte are employees of Drug Target ID, Ltd., but their activities at this company do not constitute competing interests with regard to this paper. The remaining authors declare no potential conflict of interest.

## Notes

### Competing Interest Statement

The authors have declared no competing interest.

### Funding Statement

In LEAP the results leading to this publication have received funding from the Innovative Medicines Initiative 2 Joint Undertaking under grant agreement No 777394 for the project AIMS-2-TRIALS. This Joint Undertaking receives support from the European Unions Horizon 2020 research and innovation programme and EFPIA and AUTISM SPEAKS Autistica SFARI. Any views expressed are those of the author(s) and not necessarily those of the funders (IHI-JU2).
In TACTICS the research leading to these results was supported by the European Communitys Seventh Framework Program (FP7/2007-2013) TACTICS under grant agreement no. 278948 the Innovative Medicines Initiative Joint Undertaking under grant agreement number 115300 (EU-AIMS) resources of which are composed of financial contribution from the European Unions Seventh Framework Programme (FP7- /2007 - 2013) and the European Federation of Pharmaceutical Industries and Associations (EFPIA) companies in kind contribution.

### Author Declarations

Data were collected with a research protocol that was approved by the local medical-ethics committees and after written informed consent from the volunteers or a legal guardian, across both LEAP and TACTICS cohorts included in this study. The TACTICS study was approved by the regional ethics committee of each site (Nijmegen and Utrecht: Commissie Mensgebonden Onderzoek Regio ArnhemNijmegen, 2013, NL nr 42004.091.12; Mannheim: Ethics committee of the Medical Faculty Mannheim, Heidelberg University, 2013, nr 213-616 N-MA; London: NRES Committee London - Camberwell St Giles, 2013, nr: 14/LO/1413). We obtain written informed consent from the parents of all children and oral assent from the children. When children are 12 years old they provide written informed assent themselves in addition to the parents. In case a participant or a parent retracts the consent, all data and samples will be withheld from further use for analysis and removed from the database. This is allowed at any point during the study. Participating families are regularly informed with a newsletter about study progress and resulting publications The LEAPS study was approved at each site: London-Central and Queen Square Health Research Authority Research Ethics Committee Radboud universitair medisch centrum Institut Waarborging Kwaliteit en Veiligheid Commissie Mensgebonden Onderzoek Regio Arnhem-Nijmegen (Radboud University Medical Centre Institute Ensuring Quality and Safety Committee on Research Involving Human Subjects Arnhem-Nijmegen) UMM Universitätsmedizin Mannheim, Medizinische Ethik Kommission II (UMM University Medical Mannheim, Medical Ethics Commission II) Universita Campus Bio-Medica De Roma Comitato Etico (University Campus Bio-Medical Ethics Committee De Roma) Centrala Etikprovningsnamnden (Central Ethical Review Board)

