## Supplementary material for "Estimating differing causal roles of glutamate and GABA genes on brain and behavior in autism"

### Supplementary materials

#### Inclusion and exclusion criteria LEAP

Inclusion criteria for the autism group were a clinical diagnosis of autism and age between 6 and 30 years. Autism characteristics were assessed using the Autism Diagnostic Observation Schedule Second Edition (ADOS-2; (Lord et al. 2000)) and the Autism Diagnostic Interview-Revised (ADI-R; (Rutter, Le Couteur, and Lord 2003)). For the neurotypical participants exclusion criterion consisted of parent- or self-report of any psychiatric disorder. Individuals who had a normative T-score of 70 or higher on the Social Responsiveness Scale Second Edition (SRS-2; (Bodfish et al. 2000)) were excluded. Some individuals in the autism and neurotypical groups had intellectual disability (ID) (autism=53, NTC=25), defined as an IQ score between 40 and 74. For further details of the recruitment of participants in this cohort see (Charman et al. 2017; Loth et al. 2017).

#### Inclusion and exclusion criteria TACTICS

The inclusion criteria across groups were IQ > 70, ability to speak and comprehend the native language of the recruitment location and being of Caucasian descent. To confirm a diagnosis in the autistic participants the ADI-R was used. Neurotypical participants were confirmed to not score in the clinical range for any DSM-IV axis I diagnoses using the Child Behavior Checklist (CBCL) and the Teacher Report Form (TRF) (Bordin IAS, Rocha MM, Paula CS, Teixeira MCTV, Achenbach TM, Rescola LA et al. 2013). For further details of the recruitment of participants in this cohort, see (Naaijen et al. 2016).

**Genotyping**

**LEAP.** Sample quality controls such as sex check (based on the X chromosome homozygosity rate or the median of the Log R ratio of the X and Y chromosomes), Mendelian errors (transmission errors within full trios) and Identity By State were performed using PLINK 1.90. Imputation of 17 million SNPs was performed using the 700k genotyped SNPs on the Michigan Imputation Server (Das et al. 2016). The HRC r1.1 2016 reference panel for a European population was used, as the majority of individuals in the LEAP cohort were from European ancestry. Only autosomes were imputed. Linkage disequilibrium-based SNP pruning was done for SNPs with a MAF > 1% and SNPs with an R2 < 0.1 in windows of 500kb were selected. This resulted in 546 participants with genotypic data (n = 304 autistic, n= 242 neurotypical).

**TACTICS.** Standard GWAS quality control procedures (including filtering based on minor allele frequency (MAF), Hardy-Weinberg equilibrium (p-value > 1x10e-6), single nucleotide polymorphism (SNP) call rate (> 95%), subject call rate (> 90%), principal component analysis) and imputation (1000 Genomes reference panel) were performed based on RICOPILI (Lam et al. 2020). The imputed data underwent additional quality control, in which SNPs with an imputation information score (INFO) lower than 0.8 and MAF lower than 0.05 were excluded. After this step, 5.139.250 SNPs across the autosomal genome were retained (no X-chromosome data available). This resulted in 106 participants with genotypic data (n = 31 autistic, n = 75 neurotypical).

**Table S1. Overview of included measures in all three cohorts**

| | LEAP | TACTICS | SSC | Parent/self report |
|---|---|---|---|---|
| **Glutamate PGS** | X | X | X | |
| **GABA PGS** | X | X | X | |
| **ADI** | X | | X | Parent |
| **ADOS** | X | | X | Self |
| **RBS** | X | X | X | Parent/self |
| **SRS** | X | CSBQ used as equivalent | X | Parent/self |
| **SSP** | X | | | Parent/self |
| **ADHD** | X | X | | Parent/self |
| **Depression** | X | | | Parent/self |
| **Anxiety** | X | | | Parent/self |
| **MRS glutamate (ACC, Striatum)** | | X | | |
| **fMRI successful inhibitory control (ACC, Striatum)** | X | X | | |
| **fMRI failed inhibitory control (ACC, Striatum)** | X | X | | |

Glutamate PGS, Glutamate polygenic score or autism; GABA PGS, GABA polygenic score for autism; ADI-social, Autism Diagnostic Interview-Revised Social domain; ADI-communication, Autism Diagnostic Interview-Revised Communication domain; ADI-repetitive, Autism Diagnostic Interview-Revised Restricted and Repetitive Behaviors domain; ADOS social, Autism Diagnostic Observation Schedule Second Edition Social affect; ADOS repetitive, Autism Diagnostic Observation Schedule Second Edition Restricted and Repetitive Behaviors; ADOS-total, Autism Diagnostic Observation Schedule Second Edition Total score; RBS-R, Repetitive Behavior Scale-Revised; SRS-2, Social Responsiveness Scale-Revised, CSBQ, Children's Social Behavior Questionnaire; SSP, Short Sensory Profile; ADHD, DSM-V ADHD Rating Scale; Depression, Beck Depression Inventory; Anxiety, Beck Anxiety Inventory.

**Table S2: Summary table of all genes in the glutamate gene-set**

| Gene name | Entrez gene ID | Chromosome | Start position | End position | strand | NSNPS |
|---|---|---|---|---|---|---|
| ABAT | 18 | 16 | 8768444 | 8878432 | + | 1013 |
| ALDH5A1 | 7915 | 6 | 24495197 | 24537435 | + | 297 |
| CALM1 | 801 | 14 | 90863327 | 90874619 | + | 47 |
| CALML5 | 51806 | 10 | 5540658 | 5541533 | - | 6 |
| CAMK4 | 814 | 5 | 110559947 | 110830584 | + | 1538 |
| DLG4 | 1742 | 17 | 7093209 | 7123369 | - | 102 |
| GAD1 | 2571 | 2 | 171673200 | 171717661 | + | 172 |
| GAD2 | 2572 | 10 | 26505236 | 26593491 | + | 579 |
| GLS | 2744 | 2 | 191745547 | 191830278 | + | 290 |
| GLUD1 | 2746 | 10 | 88809959 | 88854776 | - | 186 |
| GLUD2 | 2747 | X | 120181462 | 120183796 | + | |
| GLUL | 2752 | 1 | 182350839 | 182361341 | - | 55 |
| GNB1 | 2782 | 1 | 1716725 | 1822552 | - | 250 |
| GNB1L | 54584 | 22 | 19775932 | 19842462 | - | 369 |
| GNB2 | 2783 | 7 | 100271363 | 100276792 | + | 19 |
| GNB3 | 2784 | 12 | 6949375 | 6956564 | + | 34 |

| | | | | | | |
|---|---|---|---|---|---|---|
| GNB5 | 10681 | 15 | 52413123 | 52483565 | - | 486 |
| GNG10 | 2790 | 9 | 114423851 | 114432526 | + | 50 |
| GNG11 | 2791 | 7 | 93551016 | 93555826 | + | 32 |
| GNG12 | 55970 | 1 | 68167149 | 68299436 | - | 702 |
| GNG13 | 51764 | 16 | 848041 | 850733 | - | 33 |
| GNG2 | 54331 | 14 | 52327022 | 52436518 | + | 794 |
| GNG3 | 2785 | 11 | 62475066 | 62476678 | + | 5 |
| GNG4 | 2786 | 1 | 235710985 | 235814054 | - | 543 |
| GNG5 | 2787 | 1 | 84964006 | 84972262 | - | 37 |
| GNG7 | 2788 | 19 | 2511218 | 2702746 | - | 1041 |
| GOT1 | 2805 | 10 | 101156627 | 101190530 | - | 146 |
| GOT1L1 | 137362 | 8 | 37791799 | 37797664 | - | 17 |
| GOT2 | 2806 | 16 | 58741035 | 58768246 | - | 229 |
| GRIA1 | 2890 | 5 | 152870084 | 153193429 | + | 1819 |
| GRIA2 | 2891 | 4 | 158141736 | 158287227 | + | 425 |
| GRIA3 | 2892 | X | 122317996 | 122624766 | + | |
| GRIA4 | 2893 | 11 | 105480800 | 105852819 | + | 1505 |
| GRID1 | 2894 | 10 | 87359312 | 88126250 | - | 4622 |

| | | | | | | |
|---|---|---|---|---|---|---|
| GRID2 | 2895 | 4 | 93225453 | 94695707 | + | 7119 |
| GRIK1 | 2897 | 21 | 30909254 | 31312282 | - | 2258 |
| GRIK2 | 2898 | 6 | 101841584 | 102517958 | + | 3720 |
| GRIK3 | 2899 | 1 | 37261128 | 37499844 | - | 963 |
| GRIK4 | 2900 | 11 | 120382465 | 120859514 | + | 2775 |
| GRIK5 | 2901 | 19 | 42502468 | 42574278 | - | 138 |
| GRIN1 | 2902 | 9 | 140033609 | 140063214 | + | 86 |
| GRIN2A | 2903 | 16 | 9847265 | 10276611 | - | 3419 |
| GRIN2B | 2904 | 12 | 13713684 | 14133022 | - | 2569 |
| GRIN2C | 2905 | 17 | 72838162 | 72856966 | - | 93 |
| GRIN2D | 2906 | 19 | 48898132 | 48948188 | + | 222 |
| GRIN3A | 116443 | 9 | 104331634 | 104500862 | - | 942 |
| GRIN3B | 116444 | 19 | 1000437 | 1009723 | + | 108 |
| GRINA | 2907 | 8 | 145064226 | 145067596 | + | 9 |
| GRIP1 | 23426 | 12 | 66741178 | 67463014 | - | 4124 |
| GRM1 | 2911 | 6 | 146286032 | 146758782 | + | 2121 |
| GRM2 | 2912 | 3 | 51741081 | 51752629 | + | 16 |
| GRM3 | 2913 | 7 | 86273230 | 86494193 | + | 1110 |

| | | | | | | |
|---|---|---|---|---|---|---|
| GRM4 | 2914 | 6 | 33989623 | 34123399 | - | 1020 |
| GRM5 | 2915 | 11 | 88237256 | 88796846 | - | 3817 |
| GRM6 | 2916 | 5 | 178405328 | 178422124 | - | 141 |
| GRM7 | 2917 | 3 | 6902802 | 7783218 | + | 5656 |
| GRM8 | 2918 | 7 | 126078652 | 126892428 | - | 4521 |
| HOMER1 | 9456 | 5 | 78669647 | 78809659 | - | 705 |
| HOMER2 | 9455 | 15 | 83517729 | 83654905 | - | 736 |
| HOMER3 | 9454 | 19 | 19040010 | 19052041 | - | 42 |
| PICK1 | 9463 | 22 | 38453262 | 38471708 | + | 92 |
| SLC17A1 | 6568 | 6 | 25783125 | 25832287 | - | 297 |
| SLC17A2 | 10246 | 6 | 25912982 | 25930954 | - | 109 |
| SLC17A6 | 57084 | 11 | 22359667 | 22401049 | + | 208 |
| SLC17A7 | 57030 | 19 | 49932655 | 49945617 | - | 39 |
| SLC17A8 | 246213 | 12 | 100750857 | 100815837 | + | 347 |
| SLC1A1 | 6505 | 9 | 4490427 | 4587469 | + | 544 |
| SLC1A2 | 6506 | 11 | 35272752 | 35441610 | - | 1155 |
| SLC1A3 | 6507 | 5 | 36606457 | 36688436 | + | 420 |
| SLC1A4 | 6509 | 2 | 65215579 | 65250999 | + | 145 |

| | | | | | | |
|---|---|---|---|---|---|---|
| SLC1A6 | 6511 | 19 | 15060845 | 15121455 | - | 503 |
| SLC1A7 | 6512 | 1 | 53552855 | 53608304 | - | 472 |
| SLC38A1 | 81539 | 12 | 46576838 | 46663208 | - | 441 |
| SUCLG2 | 8801 | 3 | 67410884 | 67705038 | - | 1963 |

All genes in the table were included in the glutamate pathway gene-set. NSNPS, number of single nucleotide polymorphisms (SNPs).

**Table S3. Summary table of all genes in the GABA gene-set.**

| Gene name | Entrez gene ID | Chromosome | Start position | End position | strand | NSNPS |
|---|---|---|---|---|---|---|
| ABAT | 18 | 16 | 8768444 | 8878432 | + | 1013 |
| ADCY1 | 107 | 7 | 45614125 | 45762715 | + | 760 |
| ADCY10 | 55811 | 1 | 167778357 | 167883608 | - | 659 |
| ADCY2 | 108 | 5 | 7396343 | 7830194 | + | 2563 |
| ADCY3 | 109 | 2 | 25042038 | 25142602 | - | 694 |
| ADCY4 | 196883 | 14 | 24787555 | 24804277 | - | 81 |
| ADCY5 | 111 | 3 | 123001143 | 123167924 | - | 858 |
| ADCY6 | 112 | 12 | 49159975 | 49182820 | - | 81 |
| ADCY7 | 113 | 16 | 50278830 | 50352046 | + | 333 |

| | | | | | | |
|---|---|---|---|---|---|---|
| ADCY8 | 114 | 8 | 131792546 | 132053012 | - | 1901 |
| ADCY9 | 115 | 16 | 4012650 | 4166186 | - | 1082 |
| ALDH5A1 | 7915 | 6 | 24495197 | 24537435 | + | 297 |
| ALDH9A1 | 223 | 1 | 165631449 | 165667900 | - | 239 |
| AP1B1 | 162 | 22 | 29723669 | 29784754 | - | 255 |
| AP1G2 | 8906 | 14 | 24028777 | 24038754 | - | 14 |
| AP2A1 | 160 | 19 | 50270180 | 50310369 | + | 165 |
| AP2A2 | 161 | 11 | 925809 | 1012245 | + | 487 |
| AP2B1 | 163 | 17 | 33913918 | 34053436 | + | 746 |
| AP2M1 | 1173 | 3 | 183892634 | 183901879 | + | 53 |
| AP2S1 | 1175 | 19 | 47341423 | 47354203 | - | 35 |
| CACNA1A | 773 | 19 | 13317256 | 13617274 | - | 1465 |
| CACNA1B | 774 | 9 | 140772241 | 141019076 | + | 880 |
| CACNA1C | 775 | 12 | 2079952 | 2807115 | + | 3692 |
| CACNA1D | 776 | 3 | 53529076 | 53847179 | + | 1844 |
| CACNA1E | 777 | 1 | 181452447 | 181775920 | + | 1671 |
| CACNA1F | 778 | X | 49061523 | 49089833 | - | |
| CACNA1G | 8913 | 17 | 48638429 | 48704835 | + | 310 |

| | | | | | | |
|---|---|---|---|---|---|---|
| CACNA1H | 8912 | 16 | 1203241 | 1271772 | + | 422 |
| CACNA1I | 8911 | 22 | 39966758 | 40085740 | + | 591 |
| CACNA1S | 779 | 1 | 201008635 | 201081694 | - | 505 |
| CACNA2D1 | 781 | 7 | 81575760 | 82073031 | - | 3150 |
| CACNA2D2 | 9254 | 3 | 50400230 | 50540892 | - | 656 |
| CACNA2D3 | 55799 | 3 | 54156620 | 55108584 | + | 5930 |
| CACNA2D4 | 93589 | 12 | 1901123 | 2027870 | - | 775 |
| CACNB1 | 782 | 17 | 37329709 | 37353956 | - | 89 |
| CACNB2 | 783 | 10 | 18429373 | 18830688 | + | 2968 |
| CACNB3 | 784 | 12 | 49208215 | 49222726 | + | 46 |
| CACNB4 | 785 | 2 | 152689285 | 152955593 | - | 1246 |
| CACNG1 | 786 | 17 | 65040652 | 65052913 | + | 56 |
| CACNG2 | 10369 | 22 | 36956916 | 37098690 | - | 720 |
| CACNG3 | 10368 | 16 | 24266874 | 24373737 | + | 675 |
| CACNG4 | 27092 | 17 | 64960980 | 65029518 | + | 432 |
| CACNG5 | 27091 | 17 | 64831235 | 64881941 | + | 373 |
| CACNG6 | 59285 | 19 | 54494403 | 54515920 | + | 115 |
| CACNG7 | 59284 | 19 | 54412704 | 54447195 | + | 105 |

| | | | | | | |
|---|---|---|---|---|---|---|
| CACNG8 | 59283 | 19 | 54466290 | 54493469 | + | 111 |
| CATSPER1 | 117144 | 11 | 65784223 | 65793988 | - | 45 |
| CATSPER2 | 117155 | 15 | 43922772 | 43941039 | - | 63 |
| CATSPER3 | 347732 | 5 | 134303596 | 134347397 | + | 207 |
| CATSPER4 | 378807 | 1 | 26517119 | 26529033 | + | 107 |
| DNM1 | 1759 | 9 | 130965634 | 131017528 | + | 223 |
| GABARAP | 11337 | 17 | 7143738 | 7145753 | - | 5 |
| GABBR1 | 2550 | 6 | 29570005 | 29600962 | - | 219 |
| GABBR2 | 9568 | 9 | 101050364 | 101471479 | - | 2637 |
| GABRA1 | 2554 | 5 | 161274197 | 161326965 | + | 283 |
| GABRA2 | 2555 | 4 | 46246470 | 46392056 | - | 727 |
| GABRA3 | 2556 | X | 151334706 | 151619831 | - | |
| GABRA4 | 2557 | 4 | 46920917 | 46996424 | - | 406 |
| GABRA5 | 2558 | 15 | 27111866 | 27194357 | + | 158 |
| GABRA6 | 2559 | 5 | 161112658 | 161129598 | + | 81 |
| GABRB1 | 2560 | 4 | 47033295 | 47432801 | + | 2058 |
| GABRB2 | 2561 | 5 | 160715426 | 160975130 | - | 1268 |
| GABRB3 | 2562 | 15 | 26788693 | 27018935 | - | 1332 |

| | | | | | | |
|---|---|---|---|---|---|---|
| GABRD | 2563 | 1 | 1950768 | 1962192 | + | 10 |
| GABRE | 2564 | X | 151121596 | 151143156 | - | |
| GABRG1 | 2565 | 4 | 46037786 | 46126082 | - | 496 |
| GABRG2 | 2566 | 5 | 161494648 | 161582545 | + | 435 |
| GABRG3 | 2567 | 15 | 27216429 | 27778373 | + | 2556 |
| GABRP | 2568 | 5 | 170210723 | 170241051 | + | 193 |
| GABRQ | 55879 | X | 151806637 | 151821825 | + | |
| GABRR1 | 2569 | 6 | 89887223 | 89941007 | - | 344 |
| GABRR2 | 2570 | 6 | 89966840 | 90025018 | - | 405 |
| GABRR3 | 200959 | 3 | 97705527 | 97754148 | - | 264 |
| GAD1 | 2571 | 2 | 171673200 | 171717661 | + | 172 |
| GAD2 | 2572 | 10 | 26505236 | 26593491 | + | 579 |
| GNA11 | 2767 | 19 | 3094408 | 3121468 | + | 144 |
| GNA12 | 2768 | 7 | 2767739 | 2883963 | - | 883 |
| GNA13 | 10672 | 17 | 63005407 | 63052920 | - | 84 |
| GNA14 | 9630 | 9 | 80037995 | 80263232 | - | 1496 |
| GNA15 | 2769 | 19 | 3136191 | 3163766 | + | 201 |
| GNAI1 | 2770 | 7 | 79764140 | 79848725 | + | 383 |

| | | | | | | |
|---|---|---|---|---|---|---|
| GNAI2 | 2771 | 3 | 50264120 | 50296786 | + | 114 |
| GNAI3 | 2773 | 1 | 110091186 | 110138465 | + | 181 |
| GNAL | 2774 | 18 | 11689014 | 11885684 | + | 1003 |
| GNAO1 | 2775 | 16 | 56225251 | 56391356 | + | 866 |
| GNAQ | 2776 | 9 | 80335189 | 80646219 | - | 1344 |
| GNAS | 2778 | 20 | 57414756 | 57486250 | + | 323 |
| GNAT1 | 2779 | 3 | 50229043 | 50235129 | + | 12 |
| GNAT2 | 2780 | 1 | 110145889 | 110155705 | - | 45 |
| GNAZ | 2781 | 22 | 23412669 | 23467224 | + | 256 |
| GNB1 | 2782 | 1 | 1716725 | 1822552 | - | 250 |
| GNB1L | 54584 | 22 | 19775932 | 19842462 | - | 369 |
| GNB2 | 2783 | 7 | 100271363 | 100276792 | + | 19 |
| GNB3 | 2784 | 12 | 6949375 | 6956564 | + | 34 |
| GNB4 | 59345 | 3 | 179113876 | 179169371 | - | 290 |
| GNB5 | 10681 | 15 | 52413123 | 52483565 | - | 486 |
| GNG10 | 2790 | 9 | 114423851 | 114432526 | + | 50 |
| GNG11 | 2791 | 7 | 93551016 | 93555826 | + | 32 |
| GNG12 | 55970 | 1 | 68167149 | 68299436 | - | 702 |

| | | | | | | |
|---|---|---|---|---|---|---|
| GNG13 | 51764 | 16 | 848041 | 850733 | - | 33 |
| GNG2 | 54331 | 14 | 52327022 | 52436518 | + | 794 |
| GNG3 | 2785 | 11 | 62475066 | 62476678 | + | 5 |
| GNG4 | 2786 | 1 | 235710985 | 235814054 | - | 543 |
| GNG5 | 2787 | 1 | 84964006 | 84972262 | - | 37 |
| GNG7 | 2788 | 19 | 2511218 | 2702746 | - | 1041 |
| GPHN | 10243 | 14 | 66974125 | 67648525 | + | 3011 |
| GPR37 | 2861 | 7 | 124385655 | 124406079 | - | 81 |
| KCNH2 | 3757 | 7 | 150642044 | 150675402 | - | 179 |
| KCNN1 | 3780 | 19 | 18062111 | 18110133 | + | 207 |
| KCNN2 | 3781 | 5 | 113698016 | 113832197 | + | 840 |
| KCNN3 | 3782 | 1 | 154669938 | 154842754 | - | 925 |
| KCNN4 | 3783 | 19 | 44270685 | 44286269 | - | 72 |
| KCNQ2 | 3785 | 20 | 62031561 | 62103993 | - | 607 |
| KCNQ3 | 3786 | 8 | 133133105 | 133493004 | - | 2095 |
| MRAS | 22808 | 3 | 138066490 | 138124377 | + | 307 |
| NSF | 4905 | 17 | 44668035 | 44834830 | + | 108 |
| OPN1SW | 611 | 7 | 128412543 | 128415844 | - | 20 |

| | | | | | | |
|---|---|---|---|---|---|---|
| RPS27A | 6233 | 2 | 55459039 | 55462989 | + | 27 |
| SLC32A1 | 140679 | 20 | 37353105 | 37358015 | + | 20 |
| SLC6A1 | 6529 | 3 | 11034420 | 11080935 | + | 267 |
| SLC6A11 | 6538 | 3 | 10857917 | 10980146 | + | 739 |
| SLC6A12 | 6539 | 12 | 299243 | 323740 | - | 169 |
| SLC6A13 | 6540 | 12 | 329787 | 372039 | - | 322 |
| UBA52 | 7311 | 19 | 18674576 | 18688270 | + | 83 |
| UBB | 7314 | 17 | 16284367 | 16286059 | + | 7 |
| UBC | 7316 | 12 | 125396192 | 125399587 | - | 23 |
| UBD | 10537 | 6 | 29523389 | 29527702 | - | 42 |
| UBQLN1 | 29979 | 9 | 86274878 | 86323168 | - | 265 |

All genes in the table were included in the GABA pathway gene-set. NSNPS, number of single nucleotide polymorphisms (SNPs).

#### Neuroimaging

##### fMRI acquisition

**LEAP.** Structural brain images were acquired on 3 T MRI scanners at all sites, with T1-weighted MPRAGE sequences at all sites, except the London site which used IR-SPGR, which were used for registration of the functional scans. Details on the structural and functional scan parameters can be found in Table S3.

**TACTICS.** Structural T1-weighted scans were acquired based on the ADNI GO protocols (Jack et al. 2008; 2010), which were used for registration of the functional scans and voxel placement for the $^{1}$H-MRS. Spectra were acquired using a point resolved spectroscopy sequence (PRESS) with a chemically selective water suppression (CHESS) (Haase et al. 1985) from the midline pregenual ACC and the left dorsal striatum covering caudate and putamen with an 8 cm$^{3}$ voxel size (2 * 2 * 2). Voxel locations were adjusted to maximize the amount of gray matter (GM) and minimize the cerebrospinal fluid (CSF) content to keep the quality of the data as high as possible. Details on the structural, functional and $^{1}$H-MRS scan parameters can be found in Table S4.

***Magnetic resonance spectroscopy.*** Glutamate concentrations were estimated using Linear Combination Model (LCModel), with water as reference (Provencher 2001; 2014). Tissue correction and partial volume effects was calculated using the formula:

$$Metabolite_{corrected} = Metabolite_{Raw} \times \left(\frac{(43\,300 \times f_{GM} + 35\,880 \times f_{WM} + 55\,556 \times f_{CSF})}{35\,880}\right) \times \left(\frac{1}{(1 - f_{CSF})}\right)$$

where 43300 is the water concentration in millimolar for gray-matter, 35880 for white-matter, and 55556 for cerebrospinal fluid (CSF), as described in the LCModel manual (Provencher 2001). Quality control criteria were the signal-to-noise ratio of ≥ 15, Cramér-Rao lower bounds ≤ 20% and FWHM ≤ 0.1 parts per million. This resulted in data available from 44 participants. Example spectra can be seen in Figure S1 and raw glutamate levels can be found in Table S5.

#### Table S4: Scanner parameters LEAP

| Sequence | Site | TR/TE/TI (ms) | Flip angle | Slices | Coverage | Thickness (mm) | Resolution (mm³) | FOV (mm) | Gap (mm) | Volumes | Matrix |
|---|---|---|---|---|---|---|---|---|---|---|---|
| T1 | Cambridge (Siemens) | 2.3/2.95/900 | 9 | 176 | | | | | | | |
| | London (GE) | 7.31/3.02/400 | 11 | 196 | | | | | | | |
| | Mannheim (Siemens) | 2.3/2.93/900 | 9 | 176 | 256*256 | 1.2 | 1.1*1.1*1.2 | 270 | | - | - |
| | Nijmegen (Siemens) | 2.3/2.93/900 | 9 | 176 | | | | | | | |
| | Rome (GE) | 5.96/1.76/900 | 11 | 172 | | | | | | | |
| | Utrecht (Philips) | 6.76/3.1/900 | 9 | 170 | | | | | | | |
| Functional MRI | All | 2/30/- | 80 | 28 | – | 4 | 3 x 3 | 192x192 | 1 | 306 (312 Nijmegen) | 64 x 64 |

Abbreviations: FA, flip angle; FOV, field of view; TE, echo time; TR, repetition time.

#### Table S5: Scanner parameters TACTICS

| Sequence | Site | TR/TE/TI (ms) | Flip angle | Field of view (mm) | Matrix RL/AP/slices | Voxel – size (mm) | Gap (%) | Parallel Imaging | Averages Water suppressed/ unsuppressed |
|---|---|---|---|---|---|---|---|---|---|
| T1 | Nijmegen (Siemens) | 2300*/2.98/900 | 9 | 256 | 212/256/176 | 1.0*1.0*1.2 | NA | 2 | NA |
| | Mannheim (Siemens) | 2300*/2.98/900 | 9 | 270 | 212/254/176 | 1.1*1.1*1.2 | NA | 2 | NA |
| | London (GE) | 7.31*/3.02/400 | 11 | 270 | 256/256/196 | 1.1*1.1*1.2 | NA | 1.75 | NA |
| $^{1}$H-MRS PRESS | All | 3000/30/- | NA | NA | NA | 20*20*20 | NA | NA | 96/16 |
| Functional MRI | All | 2070/35/- | 74 | 192 | 192/192/36 | 3.0*3.0*3.0 | 13 | 2 | NA |

*As provided by the manufacturer. GE defines a TR as the time between excitation pulses, while Siemens defines TR as the time between inversion recovery pulses.

#### Figure S1: [1]H-MRS voxel placement in TACTICS cohort

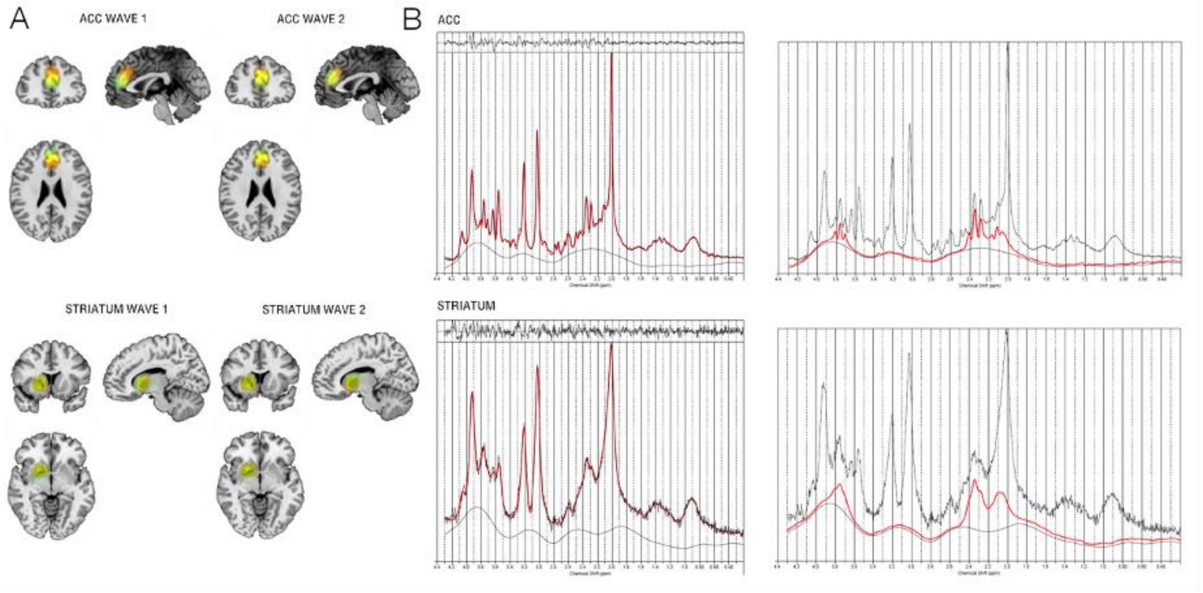


A: Superposition on the MNI152 template of all individual voxel placements in ACC and striatum, for ASD (red), OCD (blue) and controls (yellow). The placements were consistent across diagnoses, as seen by the large overlap of voxels. B: Example spectra of a 3T proton magnetic resonance spectroscopy (1H-MRS) Linear Combination (LC) Model spectral fit in ACC and striatum from one of the control participants. The top of the images represents the residuals. The black line represents frequency-domain data, the red line is the LCModel fit. The right images show the fits for glutamate only.

**fMRI preprocessing**

The LEAP data were preprocessed using the Statistical Parametric Mapping software (SPM12; http://www.fil.ion.ucl.ac.uk/spm/software/spm12/). Acquisition time correction was followed by a two-step realignment procedure to the mean functional image, coregistration of the functional data to the individual anatomical scan, followed by unified segmentation and normalization to standard stereotactic space as defined by the Montreal Neurological Institute (MNI), and smoothing with a 8mm full-width-at-half-maximum (FWHM) Gaussian Kernel. For a subset of participants from Mannheim, preprocessing additionally included bias correction of the mean image during coregistration, to adjust for measurements performed without prescan normalize option.

The TACTICS data were preprocessed using FSL (https://fsl.fmrib.ox.ac.uk/fsl/). Head movement was corrected by realigning to the middle volume (MCFLIRT; (Jenkinson et al. 2002). Grand mean scaling and spatial smoothing was done with a Gaussian kernel at FWHM of 6 mm. ICA-AROMA was used to remove secondary-head motion, followed by nuisance regression to remove CSF and white matter signal, and high-pass filtering (100 s). The fMRI data was coregistered to each participant's anatomical scan using boundary-based registration by non-linear registration FSL-FNIRT (Andersson, Jenkinson, and Smith 2007). Lastly, coregistration to the MNI template was done using a 6 mm FWHM.

**fMRI inhibitory control tasks**

In the LEAP cohort the inhibitory control task is a modified version of a combined flanker-go/nogo task (Meyer-Lindenberg et al. 2006), where participants are asked to press a left or right button depending on the direction of an arrow presented at the center of the screen. This arrow is flanked by arrows pointing either in the same direction (congruent), opposite direction (incongruent) or flanked by x's (neutral) to the centrally presented arrow. If the arrow is flanked by x's the participant was asked to withhold a response (no-go). In the TACTICS cohort the inhibitory control task consists of a stop-signal task (Rubia et al. 2003), where one arrow is presented on a screen and participants are asked to press a left or right button depending on the direction of the arrow presented on the screen. In 20% of trials the arrow is followed by a stop cue (arrow pointing upwards) and the participant is asked to withhold a response. The time between the stimulus and stop-signal (stop-signal delay, SSD) is adaptive depending on the participants performance, ensuring successful inhibition in approximately 50% of stop-trials.

In the LEAP cohort, successful inhibitory control is defined as nogo trials - failed trials, failed inhibitory control is defined as failed trials - congruent or neutral trials. Please note that failed trials comprise all committed errors including omission errors to nogo trials, interference errors to incongruent trials and omission errors to congruent, incongruent and neutral trials. In the TACTICS cohort successful inhibitory control is defined as successful stop trials - failed stop, and failed inhibitory control is defined as failed stop - successful go trials. The second level analyses of these contrasts use full-factorial designs where t-contrasts are applied to the first level contrast maps. These contrasts are created to capture inhibitory control mechanisms as similarly as possible across the cohorts.

#### Table S6. LEAP demographics

| | N | Neurotypical (N=253) | | Autism (N=343) | | Test statistic | df | p-value |
|---|---|---|---|---|---|---|---|---|
| **Sex, m/f** | | 163/90 | | 244/99 | | t = -1.73 | 525.71 | 0.08 |
| | **N** | **Mean** | **SD** | **Mean** | **SD** | | | |
| **Age** | | 17.49 | 5.84 | 17.35 | 5.49 | t = -0.31 | 523.76 | 0.75 |
| **IQ** | | 105.48 | 19.96 | 99.05 | 17.27 | t = -4.21 | 679.04 | < 0.001 |
| **SRS-2** | 546 | 28.58 | 23.17 | 89.11 | 30.81 | t = 26.20 | 543.12 | < 0.001 |
| **RBS-R** | 432 | 2.53 | 8.43 | 16.36 | 13.96 | t = 12.79 | 416.78 | < 0.001 |
| **SSP** | 323 | 176.94 | 15.62 | 139.43 | 27.27 | t = -15.72 | 320.46 | < 0.001 |
| **ADI-R** Social | 335 | - | - | 16.67 | 6.69 | - | - | - |
| Communication | 335 | - | - | 13.27 | 5.59 | - | - | - |
| Restricted repetitive | 335 | - | - | 4.27 | 2.65 | - | - | - |
| **ADOS-2** | | | | | | - | - | - |
| Social affect | 336 | - | - | 6.19 | 2.58 | | | |
| Restrictive | | | | | | - | - | - |
| repetitive | 336 | - | - | 4.65 | 2.69 | | | |

SD, standard deviation; df, degrees of freedom; SRS-2, Social Responsiveness Scale 2nd edition; RBS-R, Repetitive Behavior Scale - Revised; SSP, Short Sensory Profile; ADI-R, Autism Diagnostic Interview-Revised; Restricted repetitive, Restrictive Repetitive Behaviors domain; Communication, ADI-R Communication domain; Social, ADI-R Social domain; ADOS-2, Autism Diagnostic Observation Schedule 2nd edition; Calibrated severity, ADOS-2 Calibrated Severity Score; Social affect, ADOS-2 Social Affect.

#### Table S7. TACTICS demographics

| | | Neurotypical (N=100) | | Autism (N=60) | | Test statistic | df | p-value |
|---|---|---|---|---|---|---|---|---|
| **Sex, m/f** | | 70/30 | | 45/15 | | t =0.69 | 129.63 | 0.49 |
| | **N** | **Mean** | **SD** | **Mean** | **SD** | | | |
| **Age** | | 10.76 | 1.24 | 10.81 | 1.52 | t= -0.20 | 105.36 | 0.84 |
| **IQ** | | 110.09 | 11.47 | 107.99 | 15.12 | t = -0.93 | 99.70 | 0.36 |
| **RBS-R** | 159 | 0.95 | 1.88 | 22.25 | 20.12 | t = 8.11 | 58.60 | < 0.0001 |
| **ADI-R** Social Communicatio | 55 | - | - | 18.24 | 5.33 | - | - | - |
| n | 56 | - | - | 13.38 | 3.73 | - | - | - |
| Restricted repetitive | 55 | - | - | 3.62 | 2.61 | - | - | - |

SD, standard deviation; df, degrees of freedom; RBS-R, Repetitive Behavior Scale - Revised; ADI-R, Autism Diagnostic Interview-Revised; Restricted repetitive, Restrictive Repetitive Behaviors domain; Communication, ADI-R Communication domain; Social, AD-RI Social domain.

#### Table S8. SSC demographics

| | | Autism (N=2756) | |
|---|---|---|---|
| **Sex, m/f** | | 2382/374 | |
| | **N** | **Mean** | **SD** |
| **Age** | | 9.03 | 3.57 |
| **IQ** | | 81.15 | 27.96 |
| **SRS-2** | 2747 | 98 | 27.01 |
| **RBS-R** | 2754 | 27.14 | 17.39 |
| **ADI-R** | | | |
| Social | 2755 | 20.34 | 5.71 |
| Communication | 2422 | 16.5 | 4.26 |
| Restricted repetitive | 2755 | 6.52 | 2.50 |
| **ADOS-2** | | | |
| Social affect | 2756 | 13.33 | 4.16 |
| Restrictive repetitive | 2756 | 3.96 | 2.06 |

SD, standard deviation; df, degrees of freedom; SRS-2, Social Responsiveness Scale 2nd edition; RBS-R, Repetitive Behavior Scale - Revised; ADI-R, Autism Diagnostic Interview-Revised; Restricted repetitive, Restrictive Repetitive Behaviors domain; Communication, ADI-R Communication domain; Social, ADI-R Social domain; ADOS-2, Autism Diagnostic Observation Schedule 2nd edition; Calibrated severity, ADOS-2 Calibrated Severity Score; Social affect, ADOS-2 Social Affect.

#### Table S9: LEAP Autistic participants, all edges

| | Sex | Age | IQ | SRS | RBS | SSP | Glu PGS | GABA PGS | ADHD | Anxiety | Depression | ACC succesful | ACC failed | Striatum sucessful | Striatum failed | ADI social | ADI comm | ADI repetitive | ADOS social | ADOS repetitive |
|---|---|---|---|---|---|---|---|---|---|---|---|---|---|---|---|---|---|---|---|---|
| Sex | 0 | 0,0669 | 0,0728 | 0,0458 | 0,043 | 0,0537 | 0,0511 | 0,0758 | 0,091 | 0,195 | 0,57 | 0,071 | 0,0526 | 0,0835 | 0,0505 | 0,0797 | 0,0648 | 0,4024 | 0,3573 | 0,4163 |
| Age | 0 | 0 | 0,1203 | 0,0413 | 0,089 | 0,0672 | 0,0492 | 0,0518 | 1 | 0,0456 | 0,0707 | 0,0545 | 0,134 | 0,1057 | 0,0583 | 0,044 | 0,318 | 0,0649 | 0,0577 | 0,1842 |
| IQ | 0 | 0 | 0 | 0,1182 | 0,1849 | 0,0519 | 0,0671 | 0,0502 | 0,2004 | 0,0452 | 0,0396 | 0,0811 | 0,0844 | 0,0733 | 0,4784 | 0,5483 | 0,0613 | 0,0711 | 0,9885 | 0,0467 |
| SRS | 0 | 0 | 0 | 0 | 1 | 0,908 | 0,0513 | 0,1002 | 1 | 0,0751 | 0,0912 | 0,0563 | 0,0631 | 0,0758 | 0,0515 | 0,968 | 0,4484 | 0,0697 | 0,5183 | 0,0761 |
| RBS | 0 | 0 | 0 | 0 | 0 | 1 | 0,0532 | 0,1113 | 0,5212 | 0,5231 | 0,0532 | 0,0863 | 0,0675 | 0,0524 | 0,0587 | 0,0963 | 0,0899 | 1 | 0,1902 | 0,2207 |
| SSP | 0 | 0 | 0 | 0 | 0 | 0 | 0,0457 | 0,3515 | 0,9998 | 0,2873 | 0,0532 | 0,0491 | 0,1687 | 0,0497 | 0,3751 | 0,1999 | 0,1793 | 0,1471 | 0,0618 | 0,0586 |
| Glu PGS | 0 | 0 | 0 | 0 | 0 | 0 | 0 | 0,9745 | 0,0507 | 0,0611 | 0,0668 | 0,0619 | 0,0584 | 0,0846 | 0,1208 | 0,1149 | 0,9547 | 0,0591 | 0,0485 | 0,2904 |
| GABA PGS | 0 | 0 | 0 | 0 | 0 | 0 | 0 | 0 | 0,1115 | 0,0746 | 0,0811 | 0,209 | 0,4843 | 0,2856 | 0,2925 | 0,0512 | 0,0446 | 0,0609 | 0,0561 | 0,238 |
| ADHD | 0 | 0 | 0 | 0 | 0 | 0 | 0 | 0 | 0 | 0,0552 | 0,9835 | 0,0631 | 0,0511 | 0,0641 | 0,0508 | 0,056 | 0,0533 | 0,0583 | 0,0489 | 0,1954 |
| Anxiety | 0 | 0 | 0 | 0 | 0 | 0 | 0 | 0 | 0 | 0 | 1 | 0,0961 | 0,3915 | 0,075 | 0,1233 | 0,0665 | 0,0642 | 0,0511 | 0,2457 | 0,0532 |
| Depression | 0 | 0 | 0 | 0 | 0 | 0 | 0 | 0 | 0 | 0 | 0 | 0,0956 | 0,1184 | 0,0879 | 0,1364 | 0,0586 | 0,066 | 0,0793 | 0,5495 | 0,0415 |
| ACC succesful | 0 | 0 | 0 | 0 | 0 | 0 | 0 | 0 | 0 | 0 | 0 | 0 | 1 | 1 | 0,1325 | 0,1044 | 0,3368 | 0,1348 | 0,1283 | 0,1531 |
| ACC failed | 0 | 0 | 0 | 0 | 0 | 0 | 0 | 0 | 0 | 0 | 0 | 0 | 0 | 0,0898 | 1 | 0,0504 | 0,0478 | 0,0705 | 0,0424 | 0,0755 |
| Striatum sucessful | 0 | 0 | 0 | 0 | 0 | 0 | 0 | 0 | 0 | 0 | 0 | 0 | 0 | 0 | 1 | 0,052 | 0,162 | 0,5411 | 0,0854 | 0,1276 |
| Striatum failed | 0 | 0 | 0 | 0 | 0 | 0 | 0 | 0 | 0 | 0 | 0 | 0 | 0 | 0 | 0 | 0,0467 | 0,0579 | 0,0753 | 0,1673 | 0,231 |
| ADI social | 0 | 0 | 0 | 0 | 0 | 0 | 0 | 0 | 0 | 0 | 0 | 0 | 0 | 0 | 0 | 0 | 1 | 0,6946 | 0,1417 | 0,0627 |
| ADI comm | 0 | 0 | 0 | 0 | 0 | 0 | 0 | 0 | 0 | 0 | 0 | 0 | 0 | 0 | 0 | 0 | 0 | 0,9999 | 0,0487 | 0,0951 |
| ADI repetitive | 0 | 0 | 0 | 0 | 0 | 0 | 0 | 0 | 0 | 0 | 0 | 0 | 0 | 0 | 0 | 0 | 0 | 0 | 0,1195 | 0,8758 |
| ADOS social | 0 | 0 | 0 | 0 | 0 | 0 | 0 | 0 | 0 | 0 | 0 | 0 | 0 | 0 | 0 | 0 | 0 | 0 | 0 | 0,9891 |
| ADOS repetitive | 0 | 0 | 0 | 0 | 0 | 0 | 0 | 0 | 0 | 0 | 0 | 0 | 0 | 0 | 0 | 0 | 0 | 0 | 0 | 0 |

Red colors indicate edges of 80% reliability and above, yellow colors indicate edges between 60-80% reliability, green colors indicate below 5% reliability. Note that numbers are rounded and there may therefore be some threshold numbers with different colors. SRS, Social Responsiveness Scale 2nd edition; RBS, Repetitive Behavior Scale - Revised; SSP, Social Responsiveness Scale-Revised; Glu PGS, Glutamate polygenic score for autism; GABA PGS, GABA polygenic score for autism; ADHD, DSM-5 ADHD-Rating Scale; Anxiety, Beck Anxiety Inventory; Depression, Beck Depression Inventory-II; ACC successful, BOLD signal in ACC during successful inhibitory control; Striatum successful, BOLD signal in striatum during successful inhibitory control; ACC failed, BOLD signal in ACC during failed inhibitory control; Striatum failed, BOLD signal in striatum during failed inhibitory control; ADI social, Autism Diagnostic Interview-Revised Social domain; ADI comm, Autism Diagnostic Interview-Revised Communication domain; ADI repetitive, Autism Diagnostic Interview-Revised Restricted and Repetitive Behaviors domain; ADOS social, Autism Diagnostic Observation Schedule Second Edition Social affect; ADOS repetitive, Autism Diagnostic Observation Schedule Second Edition Restricted and Repetitive Behaviors.

#### Table S10: LEAP Autistic participants, all correlations

| | Sex | Age | IQ | SRS | RBS | SSP | Glu PGS | GABA PGS | ADHD | Anxiety | Depression | ACC succesful | ACC failed | Striatum sucessful | Striatum failed | ADI social | ADI comm | ADI repetitive | ADOS social | ADOS repetitive |
|---|---|---|---|---|---|---|---|---|---|---|---|---|---|---|---|---|---|---|---|---|
| Sex | 1 | -0,043 | -0,035 | 0,0086 | -0,039 | 0,0215 | 0,0146 | 0,0426 | -0,075 | 0,1807 | 0,1932 | 0,015 | 0,0634 | 0,0619 | 0,0008 | -0,088 | -0,076 | -0,148 | -0,166 | -0,155 |
| Age | -0,043 | 1 | -0,065 | -0,193 | -0,236 | 0,2283 | 0,0478 | -0,021 | -0,347 | -0,074 | -0,064 | 0,0244 | -0,051 | -0,032 | 0,0217 | -0,084 | -0,181 | -0,03 | 0,0306 | 0,0873 |
| IQ | -0,035 | -0,065 | 1 | -0,224 | -0,223 | 0,0652 | -0,029 | -0,025 | -0,203 | -0,067 | -0,042 | 0,0329 | 0,0661 | -0,006 | 0,1327 | -0,211 | -0,125 | -0,004 | -0,239 | -0,079 |
| SRS | 0,0086 | -0,193 | -0,224 | 1 | 0,701 | -0,598 | -0,018 | 0,1007 | 0,5989 | 0,292 | 0,3019 | -0,065 | 0,0015 | -0,065 | -0,041 | 0,4168 | 0,3857 | 0,3203 | 0,2047 | 0,1574 |
| RBS | -0,039 | -0,236 | -0,223 | 0,701 | 1 | -0,662 | -0,023 | 0,1056 | 0,5507 | 0,2934 | 0,2009 | -0,069 | -0,061 | -0,047 | -0,055 | 0,3619 | 0,3295 | 0,4292 | 0,1844 | 0,1946 |
| SSP | 0,0215 | 0,2283 | 0,0652 | -0,598 | -0,662 | 1 | 0,0559 | -0,152 | -0,558 | -0,274 | -0,186 | -0,023 | 0,1381 | -0,006 | 0,152 | -0,344 | -0,331 | -0,34 | -0,095 | -0,124 |
| Glu PGS | 0,0146 | 0,0478 | -0,029 | -0,018 | -0,023 | 0,0559 | 1 | 0,1903 | 0,0006 | -0,011 | 0,0082 | -0,025 | 0,0201 | -0,05 | 0,0721 | -0,066 | -0,193 | -0,089 | -0,015 | -0,12 |
| GABA PGS | 0,0426 | -0,021 | -0,025 | 0,1007 | 0,1056 | -0,152 | 0,1903 | 1 | 0,1077 | 0,0747 | 0,0633 | 0,1725 | -0,193 | 0,1752 | -0,185 | 0,0425 | -0,006 | 0,0611 | 0,0441 | 0,1143 |
| ADHD | -0,075 | -0,347 | -0,203 | 0,5989 | 0,5507 | -0,558 | 0,0006 | 0,1077 | 1 | 0,2228 | 0,3227 | -0,035 | 0,0261 | -0,048 | -0,008 | 0,2077 | 0,2246 | 0,2079 | 0,0115 | 0,1647 |
| Anxiety | 0,1807 | -0,074 | -0,067 | 0,292 | 0,2934 | -0,274 | -0,011 | 0,0747 | 0,2228 | 1 | 0,688 | -0,062 | 0,1951 | -0,048 | 0,164 | 0,0344 | 0,0449 | -0,009 | -0,184 | 0,0083 |
| Depression | 0,1932 | -0,064 | -0,042 | 0,3019 | 0,2009 | -0,186 | 0,0082 | 0,0633 | 0,3227 | 0,688 | 1 | -0,149 | 0,1986 | -0,129 | 0,1741 | -0,041 | -0,051 | -0,08 | -0,199 | -0,048 |
| ACC succesful | 0,015 | 0,0244 | 0,0329 | -0,065 | -0,069 | -0,023 | -0,025 | 0,1725 | -0,035 | -0,062 | -0,149 | 1 | -0,63 | 0,7978 | -0,509 | -0,085 | -0,124 | -0,104 | -0,068 | 0,0995 |
| ACC failed | 0,0634 | -0,051 | 0,0661 | 0,0015 | -0,061 | 0,1381 | 0,0201 | -0,193 | 0,0261 | 0,1951 | 0,1986 | -0,63 | 1 | -0,455 | 0,7256 | -0,046 | -0,014 | 0,0225 | -0,068 | -0,067 |
| Striatum sucessful | 0,0619 | -0,032 | -0,006 | -0,065 | -0,047 | -0,006 | -0,05 | 0,1752 | -0,048 | -0,048 | -0,129 | 0,7978 | -0,455 | 1 | -0,606 | -0,03 | -0,116 | -0,143 | -0,01 | 0,097 |
| Striatum failed | 0,0008 | 0,0217 | 0,1327 | -0,041 | -0,055 | 0,152 | 0,0721 | -0,185 | -0,008 | 0,164 | 0,1741 | -0,509 | 0,7256 | -0,606 | 1 | -0,032 | 0,0231 | 0,0549 | -0,109 | -0,126 |
| ADI social | -0,088 | -0,084 | -0,211 | 0,4168 | 0,3619 | -0,344 | -0,066 | 0,0425 | 0,2077 | 0,0344 | -0,041 | -0,085 | -0,046 | -0,03 | -0,032 | 1 | 0,6465 | 0,4053 | 0,1625 | 0,1378 |
| ADI comm | -0,076 | -0,181 | -0,125 | 0,3857 | 0,3295 | -0,331 | -0,193 | -0,006 | 0,2246 | 0,0449 | -0,051 | -0,124 | -0,014 | -0,116 | 0,0231 | 0,6465 | 1 | 0,4534 | 0,1164 | 0,1572 |
| ADI repetitive | -0,148 | -0,03 | -0,004 | 0,3203 | 0,4292 | -0,34 | -0,089 | 0,0611 | 0,2079 | -0,009 | -0,08 | -0,104 | 0,0225 | -0,143 | 0,0549 | 0,4053 | 0,4534 | 1 | 0,1511 | 0,2245 |
| ADOS social | -0,166 | 0,0306 | -0,239 | 0,2047 | 0,1844 | -0,095 | -0,015 | 0,0441 | 0,0115 | -0,184 | -0,199 | -0,068 | -0,068 | -0,01 | -0,109 | 0,1625 | 0,1164 | 0,1511 | 1 | 0,2368 |
| ADOS repetitive | -0,155 | 0,0873 | -0,079 | 0,1574 | 0,1946 | -0,124 | -0,12 | 0,1143 | 0,1647 | 0,0083 | -0,048 | 0,0995 | -0,067 | 0,097 | -0,126 | 0,1378 | 0,1572 | 0,2245 | 0,2368 | 1 |

Red colors indicate correlations 0.5 and above, yellow colors indicate correlations between 0.3-0.49, green colors indicate negative correlations from -0.3. SRS, Social Responsiveness Scale 2nd edition; RBS, Repetitive Behavior Scale - Revised; SSP, Social Responsiveness Scale-Revised; Glu PGS, Glutamate polygenic score for autism; GABA PGS, GABA polygenic score for autism; ADHD, DSM-5 ADHD-Rating Scale; Anxiety, Beck Anxiety Inventory; Depression, Beck Depression Inventory-II; ACC successful, BOLD signal in ACC during successful inhibitory control; Striatum successful, BOLD signal in striatum during successful inhibitory control; ACC failed, BOLD signal in ACC during failed inhibitory control; Striatum failed, BOLD signal in striatum during failed inhibitory control; ADI social, Autism Diagnostic Interview-Revised Social domain; ADI comm, Autism Diagnostic Interview-Revised Communication domain; ADI repetitive, Autism Diagnostic Interview-Revised Restricted and Repetitive Behaviors domain; ADOS social, Autism Diagnostic Observation Schedule Second Edition Social affect; ADOS repetitive, Autism Diagnostic Observation Schedule Second Edition Restricted and Repetitive Behaviors.

### Table S11: LEAP Neurotypical participants, all edges

| | Sex | Age | IQ | SRS | RBS | SSP | Glu PGS | GABA PGS | ADHD | Anxiety | Depression | ACC succesful | ACC failed | Striatum sucessful | Striatum failed |
|---|---|---|---|---|---|---|---|---|---|---|---|---|---|---|---|
| Sex | 0 | 0,0577 | 0,0703 | 0,2163 | 0,0875 | 0,0621 | 0,1657 | 0,1483 | 0,1187 | 0,4384 | 0,1684 | 0,1424 | 0,176 | 0,0851 | 0,1087 |
| Age | 0 | 0 | 0,0546 | 0,999 | 0,0541 | 0,9132 | 0,0836 | 0,2097 | 0,0581 | 0,9875 | 0,0727 | 0,0761 | 0,0857 | 0,0637 | 0,0655 |
| IQ | 0 | 0 | 0 | 0,9994 | 0,0719 | 0,2514 | 0,1983 | 0,0975 | 0,9486 | 0,0869 | 0,0511 | 0,0763 | 0,0957 | 0,0734 | 0,9315 |
| SRS | 0 | 0 | 0 | 0 | 0,9999 | 0,166 | 0,0762 | 0,0903 | 0,9991 | 0,0624 | 0,203 | 0,0717 | 0,063 | 0,0836 | 0,063 |
| RBS | 0 | 0 | 0 | 0 | 0 | 1 | 0,2602 | 0,0631 | 0,9598 | 0,7824 | 0,11 | 0,0734 | 0,1732 | 0,067 | 0,0609 |
| SSP | 0 | 0 | 0 | 0 | 0 | 0 | 0,0825 | 0,0944 | 0,9687 | 0,1232 | 0,2234 | 0,7796 | 0,4046 | 0,0826 | 0,083 |
| Glu PGS | 0 | 0 | 0 | 0 | 0 | 0 | 0 | 0,119 | 0,0663 | 0,0964 | 0,1317 | 0,1864 | 0,1683 | 0,0772 | 0,0844 |
| GABA PGS | 0 | 0 | 0 | 0 | 0 | 0 | 0 | 0 | 0,2291 | 0,087 | 0,0879 | 0,0917 | 0,0748 | 0,1001 | 0,0832 |
| ADHD | 0 | 0 | 0 | 0 | 0 | 0 | 0 | 0 | 0 | 0,093 | 0,9184 | 0,0947 | 0,08 | 0,118 | 0,0573 |
| Anxiety | 0 | 0 | 0 | 0 | 0 | 0 | 0 | 0 | 0 | 0 | 1 | 0,2065 | 0,0925 | 0,1523 | 0,1148 |
| Depression | 0 | 0 | 0 | 0 | 0 | 0 | 0 | 0 | 0 | 0 | 0 | 0,0795 | 0,0705 | 0,1477 | 0,1155 |
| ACC succesful | 0 | 0 | 0 | 0 | 0 | 0 | 0 | 0 | 0 | 0 | 0 | 0 | 1 | 1 | 0,1084 |
| ACC failed | 0 | 0 | 0 | 0 | 0 | 0 | 0 | 0 | 0 | 0 | 0 | 0 | 0 | 0,1047 | 1 |
| Striatum sucessful | 0 | 0 | 0 | 0 | 0 | 0 | 0 | 0 | 0 | 0 | 0 | 0 | 0 | 0 | 1 |
| Striatum failed | 0 | 0 | 0 | 0 | 0 | 0 | 0 | 0 | 0 | 0 | 0 | 0 | 0 | 0 | 0 |

Red colors indicate edges of 80% reliability and above, yellow colors indicate edges between 60-80% reliability, green colors indicate below 5% reliability. Note that numbers are rounded and there may therefore be some threshold numbers with different colors. SRS, Social Responsiveness Scale 2nd edition; RBS, Repetitive Behavior Scale - Revised; SSP, Social Responsiveness Scale-Revised; Glu PGS, Glutamate polygenic score for autism; GABA PGS, GABA polygenic score for autism; ADHD, DSM-5 ADHD-Rating Scale; Anxiety, Beck Anxiety Inventory; Depression, Beck Depression Inventory-II; ACC successful, BOLD signal in ACC during successful inhibitory control; ACC failed, BOLD signal in ACC during failed inhibitory control; Striatum successful, BOLD signal in striatum during successful inhibitory control; Striatum failed, BOLD signal in striatum during failed inhibitory control.

#### Table S12: LEAP Neurotypical participants, all correlations

| | Sex | Age | IQ | SRS | RBS | SSP | Glu PGS | GABA PGS | ADHD | Anxiety | Depression | ACC succesful | ACC failed | Striatum sucessful | Striatum failed |
|---|---|---|---|---|---|---|---|---|---|---|---|---|---|---|---|
| Sex | 1 | -0,064 | 0,0537 | -0,113 | 0,0807 | 0,0149 | -0,091 | 0,082 | -0,085 | 0,1507 | 0,1204 | -0,041 | 0,1015 | 0,0468 | 0,0773 |
| Age | -0,064 | 1 | -0,08 | 0,2716 | -0,006 | 0,2542 | -0,045 | -0,11 | 0,0865 | -0,282 | -0,161 | -0,013 | 0,1467 | 0,0015 | 0,0743 |
| IQ | 0,0537 | -0,08 | 1 | -0,43 | -0,273 | 0,3307 | 0,0924 | 0,0493 | -0,397 | -0,042 | -0,147 | -0,17 | 0,2121 | -0,115 | 0,2709 |
| SRS | -0,113 | 0,2716 | -0,43 | 1 | 0,4815 | -0,369 | 0,0371 | -0,057 | 0,4866 | 0,1798 | 0,2885 | 0,1012 | -0,091 | 0,1044 | -0,135 |
| RBS | 0,0807 | -0,006 | -0,273 | 0,4815 | 1 | -0,498 | -0,11 | 0,0048 | 0,4524 | 0,3308 | 0,3313 | 0,0567 | -0,061 | 0,0334 | -0,104 |
| SSP | 0,0149 | 0,2542 | 0,3307 | -0,369 | -0,498 | 1 | 0,0703 | -0,067 | -0,429 | -0,263 | -0,316 | -0,369 | 0,3617 | -0,281 | 0,2225 |
| Glu PGS | -0,091 | -0,045 | 0,0924 | 0,0371 | -0,11 | 0,0703 | 1 | 0,067 | -0,012 | 0,053 | -0,09 | -0,101 | 0,1043 | -0,037 | 0,0348 |
| GABA PGS | 0,082 | -0,11 | 0,0493 | -0,057 | 0,0048 | -0,067 | 0,067 | 1 | 0,1089 | -0,037 | 0,0636 | 0,0525 | -0,043 | 0,0602 | -0,045 |
| ADHD | -0,085 | 0,0865 | -0,397 | 0,4866 | 0,4524 | -0,429 | -0,012 | 0,1089 | 1 | 0,2112 | 0,3392 | 0,0892 | -0,078 | 0,082 | -0,113 |
| Anxiety | 0,1507 | -0,282 | -0,042 | 0,1798 | 0,3308 | -0,263 | 0,053 | -0,037 | 0,2112 | 1 | 0,674 | -0,016 | 0,0301 | 0,0937 | -0,051 |
| Depression | 0,1204 | -0,161 | -0,147 | 0,2885 | 0,3313 | -0,316 | -0,09 | 0,0636 | 0,3392 | 0,674 | 1 | 0,0598 | -0,014 | 0,1054 | -0,064 |
| ACC succesful | -0,041 | -0,013 | -0,17 | 0,1012 | 0,0567 | -0,369 | -0,101 | 0,0525 | 0,0892 | -0,016 | 0,0598 | 1 | -0,663 | 0,7532 | -0,502 |
| ACC failed | 0,1015 | 0,1467 | 0,2121 | -0,091 | -0,061 | 0,3617 | 0,1043 | -0,043 | -0,078 | 0,0301 | -0,014 | -0,663 | 1 | -0,453 | 0,6954 |
| Striatum sucessful | 0,0468 | 0,0015 | -0,115 | 0,1044 | 0,0334 | -0,281 | -0,037 | 0,0602 | 0,082 | 0,0937 | 0,1054 | 0,7532 | -0,453 | 1 | -0,663 |
| Striatum failed | 0,0773 | 0,0743 | 0,2709 | -0,135 | -0,104 | 0,2225 | 0,0348 | -0,045 | -0,113 | -0,051 | -0,064 | -0,502 | 0,6954 | -0,663 | 1 |

Red colors indicate correlations 0.5 and above, yellow colors indicate correlations between 0.3-0.49, green colors indicate negative correlations from -0.3. SRS, Social Responsiveness Scale 2nd edition; RBS, Repetitive Behavior Scale - Revised; SSP, Social Responsiveness Scale-Revised; Glu PGS, Glutamate polygenic score for autism; GABA PGS, GABA polygenic score for autism; ADHD, DSM-5 ADHD-Rating Scale; Anxiety, Beck Anxiety Inventory; Depression, Beck Depression Inventory-II; ACC successful, BOLD signal in ACC during successful inhibitory control; ACC failed, BOLD signal in ACC during failed inhibitory control; Striatum successful, BOLD signal in striatum during successful inhibitory control; Striatum failed, BOLD signal in striatum during failed inhibitory control.

#### Table S13: LEAP All participants, all edges

| | Diagnosis | Sex | Age | IQ | SRS | RBS | SSP | Glu PGS | GABA PGS | ADHD | Anxiety | Depression | ACC succesful | ACC failed | Striatum sucessful | Striatum failed |
|---|---|---|---|---|---|---|---|---|---|---|---|---|---|---|---|---|
| Diagnosis | 0 | 0,1228 | 0,0459 | 0,3654 | 1 | 1 | 0,0906 | 0,078 | 0,0556 | 0,0575 | 0,2075 | 0,451 | 0,1716 | 0,058 | 0,1005 | 0,0567 |
| Sex | 0 | 0 | 0,0488 | 0,0391 | 0,1852 | 0,1055 | 0,0655 | 0,0581 | 0,1339 | 0,2995 | 0,3105 | 0,2333 | 0,0551 | 0,1273 | 0,1592 | 0,0632 |
| Age | 0 | 0 | 0 | 0,131 | 0,08 | 0,0504 | 0,4689 | 0,0407 | 0,0489 | 0,2586 | 0,5424 | 0,0464 | 0,1626 | 0,0345 | 0,0551 | 0,0536 |
| IQ | 0 | 0 | 0 | 0 | 0,8831 | 0,0857 | 0,0493 | 0,0442 | 0,0364 | 0,9688 | 0,0358 | 0,0351 | 0,0481 | 0,0468 | 0,0564 | 0,9755 |
| SRS | 0 | 0 | 0 | 0 | 0 | 1 | 0,9433 | 0,0762 | 0,0624 | 1 | 0,095 | 0,9217 | 0,0843 | 0,0534 | 0,0713 | 0,0584 |
| RBS | 0 | 0 | 0 | 0 | 0 | 0 | 1 | 0,189 | 0,0709 | 0,8518 | 0,8571 | 0,059 | 0,0726 | 0,0654 | 0,0512 | 0,0544 |
| SSP | 0 | 0 | 0 | 0 | 0 | 0 | 0 | 0,201 | 0,4374 | 1 | 0,6939 | 0,0648 | 0,053 | 0,3753 | 0,05 | 0,1886 |
| Glu PGS | 0 | 0 | 0 | 0 | 0 | 0 | 0 | 0 | 0,9074 | 0,0632 | 0,0482 | 0,1346 | 0,1058 | 0,0911 | 0,0604 | 0,0679 |
| GABA PGS | 0 | 0 | 0 | 0 | 0 | 0 | 0 | 0 | 0 | 0,1189 | 0,0403 | 0,0595 | 0,0818 | 0,2913 | 0,1491 | 0,2747 |
| ADHD | 0 | 0 | 0 | 0 | 0 | 0 | 0 | 0 | 0 | 0 | 0,0657 | 1 | 0,0502 | 0,0427 | 0,0546 | 0,0439 |
| Anxiety | 0 | 0 | 0 | 0 | 0 | 0 | 0 | 0 | 0 | 0 | 0 | 1 | 0,0755 | 0,2376 | 0,1633 | 0,0751 |
| Depression | 0 | 0 | 0 | 0 | 0 | 0 | 0 | 0 | 0 | 0 | 0 | 0 | 0,0531 | 0,1009 | 0,0536 | 0,0671 |
| ACC succesful | 0 | 0 | 0 | 0 | 0 | 0 | 0 | 0 | 0 | 0 | 0 | 0 | 0 | 1 | 1 | 0,0718 |
| ACC failed | 0 | 0 | 0 | 0 | 0 | 0 | 0 | 0 | 0 | 0 | 0 | 0 | 0 | 0 | 0,0561 | 1 |
| Striatum sucessful | 0 | 0 | 0 | 0 | 0 | 0 | 0 | 0 | 0 | 0 | 0 | 0 | 0 | 0 | 0 | 1 |
| Striatum failed | 0 | 0 | 0 | 0 | 0 | 0 | 0 | 0 | 0 | 0 | 0 | 0 | 0 | 0 | 0 | 0 |

Red colors indicate edges of 80% reliability and above, yellow colors indicate edges between 60-80% reliability, green colors indicate below 5% reliability. Note that numbers are rounded and there may therefore be some threshold numbers with different colors. SRS, Social Responsiveness Scale 2nd edition; RBS, Repetitive Behavior Scale - Revised; SSP, Social Responsiveness Scale-Revised; Glu PGS, Glutamate polygenic score for autism; GABA PGS, GABA polygenic score for autism; ADHD, DSM-5 ADHD-Rating Scale; Anxiety, Beck Anxiety Inventory; Depression, Beck Depression Inventory-II; ACC successful, BOLD signal in ACC during successful inhibitory control; ACC failed, BOLD signal in ACC during failed inhibitory control; Striatum successful, BOLD signal in striatum during successful inhibitory control; Striatum failed, BOLD signal in striatum during failed inhibitory control.

#### Table S14: LEAP All participants, all correlations

| | Diagnosis | Sex | Age | IQ | SRS | RBS | SSP | Glu PGS | GABA PGS | ADHD | Anxiety | Depression | ACC succesful | ACC failed | Striatum sucessful | Striatum failed |
|---|---|---|---|---|---|---|---|---|---|---|---|---|---|---|---|---|
| Diagnosis | 1 | -0,081 | -0,009 | -0,143 | 0,7328 | 0,7009 | -0,564 | -0,056 | 0,0081 | 0,5127 | 0,4036 | 0,4428 | 0,0831 | -0,062 | 0,0686 | -0,066 |
| Sex | -0,081 | 1 | -0,047 | 0,0152 | -0,09 | -0,077 | 0,065 | -0,028 | 0,0617 | -0,1 | 0,1128 | 0,1008 | -0,028 | 0,088 | 0,0383 | 0,0401 |
| Age | -0,009 | -0,047 | 1 | -0,06 | 0,0161 | -0,087 | 0,1555 | 0,0148 | -0,042 | -0,145 | -0,153 | -0,099 | 0,0382 | 0,0359 | -1E-03 | 0,0583 |
| IQ | -0,143 | 0,0152 | -0,06 | 1 | -0,332 | -0,302 | 0,2103 | 0,0182 | -0,011 | -0,333 | -0,115 | -0,156 | -0,058 | 0,1174 | -0,037 | 0,1699 |
| SRS | 0,7328 | -0,09 | 0,0161 | -0,332 | 1 | 0,8234 | -0,719 | -0,039 | 0,025 | 0,7045 | 0,4449 | 0,5129 | 0,0628 | -0,064 | 0,0542 | -0,086 |
| RBS | 0,7009 | -0,077 | -0,087 | -0,302 | 0,8234 | 1 | -0,777 | -0,079 | 0,0503 | 0,687 | 0,4624 | 0,4652 | 0,0123 | -0,065 | 0,0197 | -0,078 |
| SSP | -0,564 | 0,065 | 0,1555 | 0,2103 | -0,719 | -0,777 | 1 | 0,0835 | -0,112 | -0,68 | -0,437 | -0,43 | -0,07 | 0,1311 | -0,05 | 0,1342 |
| Glu PGS | -0,056 | -0,028 | 0,0148 | 0,0182 | -0,039 | -0,079 | 0,0835 | 1 | 0,1308 | -0,042 | -0,008 | -0,063 | -0,056 | 0,0553 | -0,032 | 0,046 |
| GABA PGS | 0,0081 | 0,0617 | -0,042 | -0,011 | 0,025 | 0,0503 | -0,112 | 0,1308 | 1 | 0,084 | 0,0158 | 0,0481 | 0,0732 | -0,105 | 0,0838 | -0,104 |
| ADHD | 0,5127 | -0,1 | -0,145 | -0,333 | 0,7045 | 0,687 | -0,68 | -0,042 | 0,084 | 1 | 0,3841 | 0,4947 | 0,0456 | -0,044 | 0,0334 | -0,071 |
| Anxiety | 0,4036 | 0,1128 | -0,153 | -0,115 | 0,4449 | 0,4624 | -0,437 | -0,008 | 0,0158 | 0,3841 | 1 | 0,7415 | -0,009 | 0,0865 | 0,0489 | 0,0224 |
| Depression | 0,4428 | 0,1008 | -0,099 | -0,156 | 0,5129 | 0,4652 | -0,43 | -0,063 | 0,0481 | 0,4947 | 0,7415 | 1 | 0,004 | 0,0667 | 0,0238 | 0,0315 |
| ACC succesful | 0,0831 | -0,028 | 0,0382 | -0,058 | 0,0628 | 0,0123 | -0,07 | -0,056 | 0,0732 | 0,0456 | -0,009 | 0,004 | 1 | -0,643 | 0,7756 | -0,501 |
| ACC failed | -0,062 | 0,088 | 0,0359 | 0,1174 | -0,064 | -0,065 | 0,1311 | 0,0553 | -0,105 | -0,044 | 0,0865 | 0,0667 | -0,643 | 1 | -0,458 | 0,7201 |
| Striatum sucessful | 0,0686 | 0,0383 | -1E-03 | -0,037 | 0,0542 | 0,0197 | -0,05 | -0,032 | 0,0838 | 0,0334 | 0,0489 | 0,0238 | 0,7756 | -0,458 | 1 | -0,631 |
| Striatum failed | -0,066 | 0,0401 | 0,0583 | 0,1699 | -0,086 | -0,078 | 0,1342 | 0,046 | -0,104 | -0,071 | 0,0224 | 0,0315 | -0,501 | 0,7201 | -0,631 | 1 |

Red colors indicate correlations 0.5 and above, yellow colors indicate correlations between 0.3-0.49, green colors indicate negative correlations from -0.3. SRS, Social Responsiveness Scale 2nd edition; RBS, Repetitive Behavior Scale - Revised; SSP, Social Responsiveness Scale-Revised; Glu PGS, Glutamate polygenic score for autism; GABA PGS, GABA polygenic score for autism; ADHD, DSM-5 ADHD-Rating Scale; Anxiety, Beck Anxiety Inventory; Depression, Beck Depression Inventory-II; ACC successful, BOLD signal in ACC during successful inhibitory control; ACC failed, BOLD signal in ACC during failed inhibitory control; Striatum successful, BOLD signal in striatum during successful inhibitory control; Striatum failed, BOLD signal in striatum during failed inhibitory control.

#### Table S15: TACTICS All participants, all edges

| | Diagnosis | Sex | Age | IQ | RBS | Glu PGS | GABA PGS | ADHD | ACC failed | Striatum failed | ACC successful | Striatum successful | Glutamate ACC | Glutamate Striatum | CSBQ |
|---|---|---|---|---|---|---|---|---|---|---|---|---|---|---|---|
| Diagnosis | 0 | 0,0952 | 0,0778 | 0,1463 | 1 | 0,1307 | 0,1401 | 0,2128 | 0,161 | 0,094 | 0,3549 | 0,0986 | 0,0768 | 0,4723 | 0,9999 |
| Sex | 0 | 0 | 0,0701 | 0,1038 | 0,1215 | 0,1339 | 0,1498 | 0,2312 | 0,3309 | 0,0747 | 0,0764 | 0,1244 | 0,9115 | 0,607 | 0,1602 |
| Age | 0 | 0 | 0 | 0,2923 | 0,1285 | 0,0925 | 0,2864 | 0,1996 | 0,2335 | 0,1316 | 0,6363 | 0,9528 | 0,4223 | 0,0728 | 0,0831 |
| IQ | 0 | 0 | 0 | 0 | 0,1441 | 0,4697 | 0,1853 | 0,0801 | 0,079 | 1 | 0,1049 | 0,0747 | 0,0779 | 0,0944 | 0,1774 |
| RBS | 0 | 0 | 0 | 0 | 0 | 0,1466 | 0,533 | 0,6643 | 0,1617 | 0,0912 | 0,1197 | 0,0868 | 0,0742 | 0,3069 | 1 |
| Glu PGS | 0 | 0 | 0 | 0 | 0 | 0 | 0,0775 | 0,2977 | 0,2957 | 0,3521 | 0,9986 | 0,9412 | 0,0872 | 0,4663 | 0,1404 |
| GABA PGS | 0 | 0 | 0 | 0 | 0 | 0 | 0 | 0,1596 | 0,1648 | 0,1214 | 0,1063 | 0,2574 | 0,9888 | 0,3208 | 0,1047 |
| ADHD | 0 | 0 | 0 | 0 | 0 | 0 | 0 | 0 | 0,0814 | 0,0756 | 0,0806 | 0,0731 | 0,1738 | 0,1418 | 0,9996 |
| ACC failed | 0 | 0 | 0 | 0 | 0 | 0 | 0 | 0 | 0 | 1 | 0,1344 | 0,0961 | 0,1986 | 0,1643 | 0,5141 |
| Striatum failed | 0 | 0 | 0 | 0 | 0 | 0 | 0 | 0 | 0 | 0 | 0,069 | 0,073 | 0,3013 | 0,26 | 0,1304 |
| ACC successful | 0 | 0 | 0 | 0 | 0 | 0 | 0 | 0 | 0 | 0 | 0 | 0,473 | 0,3709 | 0,0799 | 0,0829 |
| Striatum successful | 0 | 0 | 0 | 0 | 0 | 0 | 0 | 0 | 0 | 0 | 0 | 0 | 0,1669 | 0,9619 | 0,0848 |
| Glutamate ACC | 0 | 0 | 0 | 0 | 0 | 0 | 0 | 0 | 0 | 0 | 0 | 0 | 0 | 0,155 | 0,1327 |
| Glutamate Striatum | 0 | 0 | 0 | 0 | 0 | 0 | 0 | 0 | 0 | 0 | 0 | 0 | 0 | 0 | 0,2536 |
| CSBQ | 0 | 0 | 0 | 0 | 0 | 0 | 0 | 0 | 0 | 0 | 0 | 0 | 0 | 0 | 0 |

Red colors indicate edges of 80% reliability and above, yellow colors indicate edges between 60-80% reliability, green colors indicate below 5% reliability. Note that numbers are rounded and there may therefore be some threshold numbers with different colors. RBS, Repetitive Behavior Scale - Revised; Glu PGS, Glutamate polygenic score for autism, GABA PGS, GABA polygenic score for autism; ADHD, Conners' Parent Rating Scale; ACC failed, BOLD signal in ACC during failed inhibitory control; Striatum failed, BOLD signal in striatum during failed inhibitory control; ACC successful, BOLD signal in ACC during successful inhibitory control; Striatum successful, BOLD signal in striatum during successful inhibitory control; Glutamate ACC, estimated glutamate concentrations in the ACC using water reference; Glutamate Striatum; Estimated glutamate concentrations in Striatum using water reference; CSBQ, Children's Social Behavior Questionnaire.

**Table S16: TACTICS All participants, all correlations**

| | Diagnosis | Sex | Age | IQ | RBS | Glu PGS | GABA PGS | ADHD | ACC failed | Striatum failed | ACC successful | Striatum successful | Glutamate ACC | Glutamate Striatum | CSBQ |
|---|---|---|---|---|---|---|---|---|---|---|---|---|---|---|---|
| Diagnosis | 1 | -0,054 | 0,0312 | -0,073 | 0,7917 | -0,111 | 0,0774 | 0,2469 | -0,158 | -0,066 | 0,1671 | 0,1089 | 0,0221 | -0,211 | 0,7959 |
| Sex | -0,054 | 1 | -0,021 | -0,051 | -0,091 | 0,1179 | 0,0945 | 0,1097 | 0,1637 | 0,0135 | -0,047 | -0,091 | -0,251 | -0,188 | -0,122 |
| Age | 0,0312 | -0,021 | 1 | -0,147 | -0,077 | 0,1383 | -0,144 | 0,1321 | 0,1342 | 0,1021 | 0,2083 | -0,269 | 0,215 | -0,001 | 0,0443 |
| IQ | -0,073 | -0,051 | -0,147 | 1 | -0,111 | 0,1764 | 0,1147 | -0,011 | 0,1439 | 0,3793 | -0,117 | -0,018 | -0,083 | -0,075 | -0,175 |
| RBS | 0,7917 | -0,091 | -0,077 | -0,111 | 1 | -0,11 | -0,052 | 0,2036 | -0,221 | -0,061 | -0,032 | -0,014 | 0,002 | -0,021 | 0,8223 |
| Glu PGS | -0,111 | 0,1179 | 0,1383 | 0,1764 | -0,11 | 1 | 0,0693 | 0,1476 | -0,204 | -0,203 | -0,396 | -0,358 | 0,0716 | -0,19 | -0,116 |
| GABA PGS | 0,0774 | 0,0945 | -0,144 | 0,1147 | -0,052 | 0,0693 | 1 | -0,111 | 0,0993 | -0,102 | -0,117 | -0,168 | 0,2945 | 0,1733 | 0,0587 |
| ADHD | 0,2469 | 0,1097 | 0,1321 | -0,011 | 0,2036 | 0,1476 | -0,111 | 1 | 0,0183 | -0,018 | -0,044 | 0,0417 | -0,116 | -0,163 | 0,3851 |
| ACC failed | -0,158 | 0,1637 | 0,1342 | 0,1439 | -0,221 | -0,204 | 0,0993 | 0,0183 | 1 | 0,5 | 0,1393 | -0,077 | -0,163 | 0,1741 | -0,279 |
| Striatum failed | -0,066 | 0,0135 | 0,1021 | 0,3793 | -0,061 | -0,203 | -0,102 | -0,018 | 0,5 | 1 | 0,0571 | 0,0122 | -0,183 | 0,1743 | -0,19 |
| ACC successful | 0,1671 | -0,047 | 0,2083 | -0,117 | -0,032 | -0,396 | -0,117 | -0,044 | 0,1393 | 0,0571 | 1 | 0,2862 | 0,175 | 0,0663 | 0,0509 |
| Striatum successful | 0,1089 | -0,091 | -0,269 | -0,018 | -0,014 | -0,358 | -0,168 | 0,0417 | -0,077 | 0,0122 | 0,2862 | 1 | -0,188 | -0,273 | 0,0773 |
| Glutamate ACC | 0,0221 | -0,251 | 0,215 | -0,083 | 0,002 | 0,0716 | 0,2945 | -0,116 | -0,163 | -0,183 | 0,175 | -0,188 | 1 | 0,1545 | -0,081 |
| Glutamate Striatum | -0,211 | -0,188 | -0,001 | -0,075 | -0,021 | -0,19 | 0,1733 | -0,163 | 0,1741 | 0,1743 | 0,0663 | -0,273 | 0,1545 | 1 | -0,195 |
| CSBQ | 0,7959 | -0,122 | 0,0443 | -0,175 | 0,8223 | -0,116 | 0,0587 | 0,3851 | -0,279 | -0,19 | 0,0509 | 0,0773 | -0,081 | -0,195 | 1 |

Red colors indicate correlations 0.5 and above, yellow colors indicate correlations between 0.3-0.49, green colors indicate negative correlations from -0.3. RBS, Repetitive Behavior Scale - Revised; Glu PGS, Glutamate polygenic score for autism, GABA PGS, GABA polygenic score for autism; ADHD, Conners' Parent Rating Scale; ACC failed, BOLD signal in ACC during failed inhibitory control; Striatum failed, BOLD signal in striatum during failed inhibitory control; ACC successful, BOLD signal in ACC during successful inhibitory control; Striatum successful, BOLD signal in striatum during successful inhibitory control; Glutamate ACC, estimated glutamate concentrations in the ACC using water reference; Glutamate Striatum; Estimated glutamate concentrations in Striatum using water reference; CSBQ, Children's Social Behavior Questionnaire.

#### Table S17: SSC All (autistic) participants, all edges

| | ADI repetitive | ADI comm | ADI social | ADOS repetitive | ADOS social | RBS | Sex | IQ | SRS | Age | Glu PRS | GABA PRS |
|---|---|---|---|---|---|---|---|---|---|---|---|---|
| ADI repetitive | 0 | 1 | 0,1185 | 0,1445 | 0,0195 | 1 | 0,804 | 0,0424 | 0,0389 | 0,02 | 0,0212 | 0,0224 |
| ADI comm | 0 | 0 | 1 | 0,582 | 0,096 | 0,0154 | 0,0144 | 0,996 | 0,025 | 0,183 | 0,1247 | 0,0276 |
| ADI social | 0 | 0 | 0 | 0,0247 | 0,9987 | 0,0263 | 0,0189 | 1 | 1 | 1 | 0,0711 | 0,0399 |
| ADOS repetitive | 0 | 0 | 0 | 0 | 1 | 0,0719 | 0,0186 | 1 | 0,0218 | 1 | 0,0399 | 0,0311 |
| ADOS social | 0 | 0 | 0 | 0 | 0 | 0,0179 | 0,0217 | 1 | 0,036 | 0,2449 | 0,0341 | 0,0236 |
| RBS | 0 | 0 | 0 | 0 | 0 | 0 | 0,0174 | 0,2614 | 1 | 0,1761 | 0,0283 | 0,0242 |
| Sex | 0 | 0 | 0 | 0 | 0 | 0 | 0 | 0 | 0,0178 | 0,0323 | 0,0491 | 0,0217 |
| IQ | 0 | 0 | 0 | 0 | 0 | 0 | 0 | 0 | 0,8759 | 0,0312 | 0,0425 | 0,0664 |
| SRS | 0 | 0 | 0 | 0 | 0 | 0 | 0 | 0 | 0 | 0,034 | 0,0244 | 0,0352 |
| Age | 0 | 0 | 0 | 0 | 0 | 0 | 0 | 0 | 0 | 0 | 0,0261 | 0,0224 |
| Glu PRS | 0 | 0 | 0 | 0 | 0 | 0 | 0 | 0 | 0 | 0 | 0 | 0,0693 |
| GABA PRS | 0 | 0 | 0 | 0 | 0 | 0 | 0 | 0 | 0 | 0 | 0 | 0 |

Red colors indicate edges of 80% reliability and above, yellow colors indicate edges between 60-80% reliability, green colors indicate below 5% reliability. Note that numbers are rounded and there may therefore be some threshold numbers with different colors. ADI repetitive, Autism Diagnostic Interview-Revised Restricted and Repetitive Behaviors domain; ADI comm, Autism Diagnostic Interview-Revised Communication domain; ADI social, Autism Diagnostic Interview-Revised Social domain; ADOS repetitive, Autism Diagnostic Observation Schedule Second Edition Restricted and Repetitive Behaviors; ADOS social, Autism Diagnostic Observation Schedule Second Edition Social affect; RBS, Repetitive Behavior Scale - Revised; SRS, Social Responsiveness Scale 2nd edition; Glu PGS, Glutamate polygenic score for autism; GABA PGS, GABA polygenic score for autism.

#### Table S18: SSC All (autistic) participants, all correlations

| | ADI repetitive | ADI comm | ADI social | ADOS repetitive | ADOS social | RBS | Sex | IQ | SRS | Age | Glu PRS | GABA PRS |
|---|---|---|---|---|---|---|---|---|---|---|---|---|
| ADI repetitive | 1 | 0,2636 | 0,2245 | 0,1321 | 0,0604 | 0,4019 | -0,061 | -0,033 | 0,2452 | 0,0592 | -0,001 | -0,003 |
| ADI comm | 0,2636 | 1 | 0,6632 | 0,2539 | 0,2632 | 0,2493 | 0,0177 | -0,34 | 0,3318 | 0,1057 | -0,042 | 0,0167 |
| ADI social | 0,2245 | 0,6632 | 1 | 0,2298 | 0,2893 | 0,2426 | 0,0184 | -0,381 | 0,4139 | 0,2384 | -0,035 | 0,0241 |
| ADOS repetitive | 0,1321 | 0,2539 | 0,2298 | 1 | 0,4568 | 0,1514 | 0,003 | -0,426 | 0,1491 | -0,211 | -0,026 | 0,0191 |
| ADOS social | 0,0604 | 0,2632 | 0,2893 | 0,4568 | 1 | 0,1084 | 0,0399 | -0,421 | 0,1679 | -0,13 | -0,023 | 0,0104 |
| RBS | 0,4019 | 0,2493 | 0,2426 | 0,1514 | 0,1084 | 1 | -0,003 | -0,177 | 0,6027 | -0,038 | -0,018 | 0,0059 |
| Sex | -0,061 | 0,0177 | 0,0184 | 0,003 | 0,0399 | -0,003 | 1 | -0,08 | 0,03 | 0,0175 | 0,0245 | 0,0053 |
| IQ | -0,033 | -0,34 | -0,381 | -0,426 | -0,421 | -0,177 | -0,08 | 1 | -0,239 | 0,0014 | 0,0271 | -0,031 |
| SRS | 0,2452 | 0,3318 | 0,4139 | 0,1491 | 0,1679 | 0,6027 | 0,03 | -0,239 | 1 | 0,1108 | -0,013 | 0,0221 |
| Age | 0,0592 | 0,1057 | 0,2384 | -0,211 | -0,13 | -0,038 | 0,0175 | 0,0014 | 0,1108 | 1 | 0,0127 | -0,006 |
| Glu PRS | -0,001 | -0,042 | -0,035 | -0,026 | -0,023 | -0,018 | 0,0245 | 0,0271 | -0,013 | 0,0127 | 1 | 0,0288 |
| GABA PRS | -0,003 | 0,0167 | 0,0241 | 0,0191 | 0,0104 | 0,0059 | 0,0053 | -0,031 | 0,0221 | -0,006 | 0,0288 | 1 |

Red colors indicate correlations 0.5 and above, yellow colors indicate correlations between 0.3-0.49, green colors indicate negative correlations from -0.3. ADI repetitive, Autism Diagnostic Interview-Revised Restricted and Repetitive Behaviors domain; ADI comm, Autism Diagnostic Interview-Revised Communication domain; ADI social, Autism Diagnostic Interview-Revised Social domain; ADOS repetitive, Autism Diagnostic Observation Schedule Second Edition Restricted and Repetitive Behaviors; ADOS social, Autism Diagnostic Observation Schedule Second Edition Social affect; RBS, Repetitive Behavior Scale - Revised; SRS, Social Responsiveness Scale 2nd edition; Glu PGS, Glutamate polygenic score for autism; GABA PGS, GABA polygenic score for autism.

#### Table S19: Post-hoc tests of differences between cohorts

| | Glutamate PGS | GABA PGS | ADI-R communication | ADI-R restricted repetitive | ADI-R social |
|---|---|---|---|---|---|
| LEAP - SSC | **t =-17.616 df = 382.02 p<2.2e-16** | **t = -7.1013 df = 375.56, p= 6.229e-12** | **t = 6.2974 df = 62.366 p = 3.408e-08** | **t = -14.728 df = 423.4 p < 2.2e-16** | **t = 3.2538 df = 60.76 p = 0.001862** |

t, t-score; df, degrees of freedom; Glutamate PGS, Glutamate polygenic score, GABA PGS, GABA polygenic score, ADI-R, Autism Diagnostic Interview-Revised; Restricted repetitive, Restrictive Repetitive Behaviors domain; Communication, ADI-R Communication domain; Social, ADI-R Social domain. LEAP, Longitudinal European Autism Project cohort, SSC, Simons Simplex Collection cohort. Significant results are marked in bold.

#### Figure S2: Polygenic scores

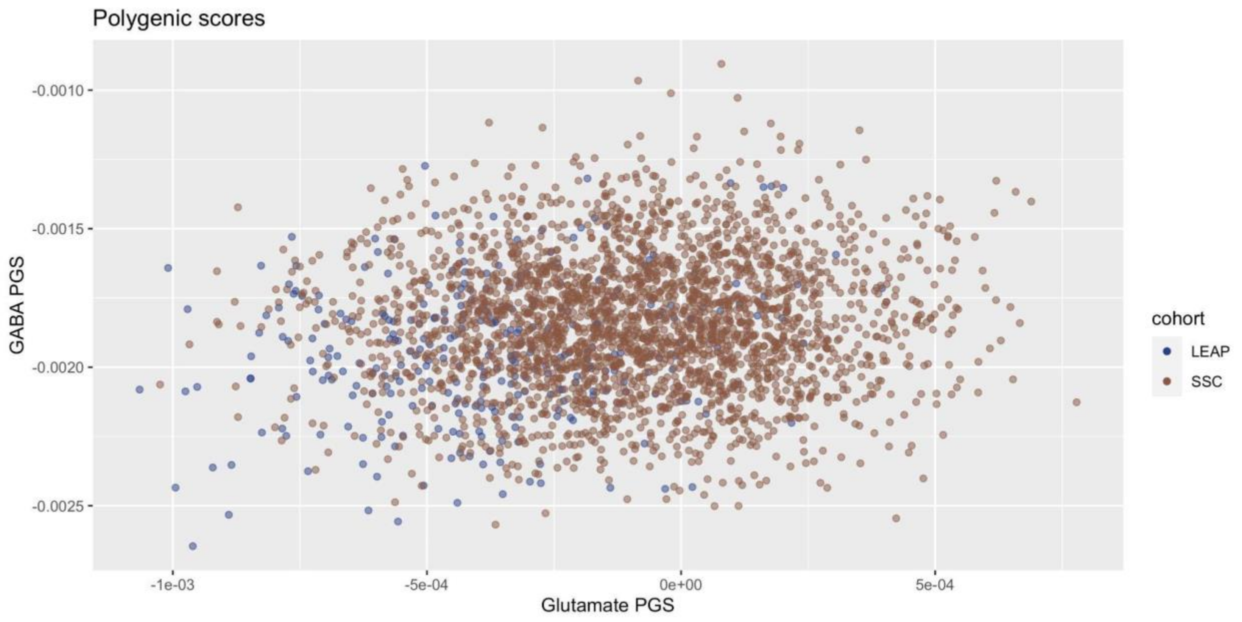


Glutamate and GABA polygenic scores in all three cohorts. SSC, Simons Simplex Collection (brown); LEAP, Longitudinal European Autism Project (blue).

#### Figure S3: ADI-R Restricted Repetitive

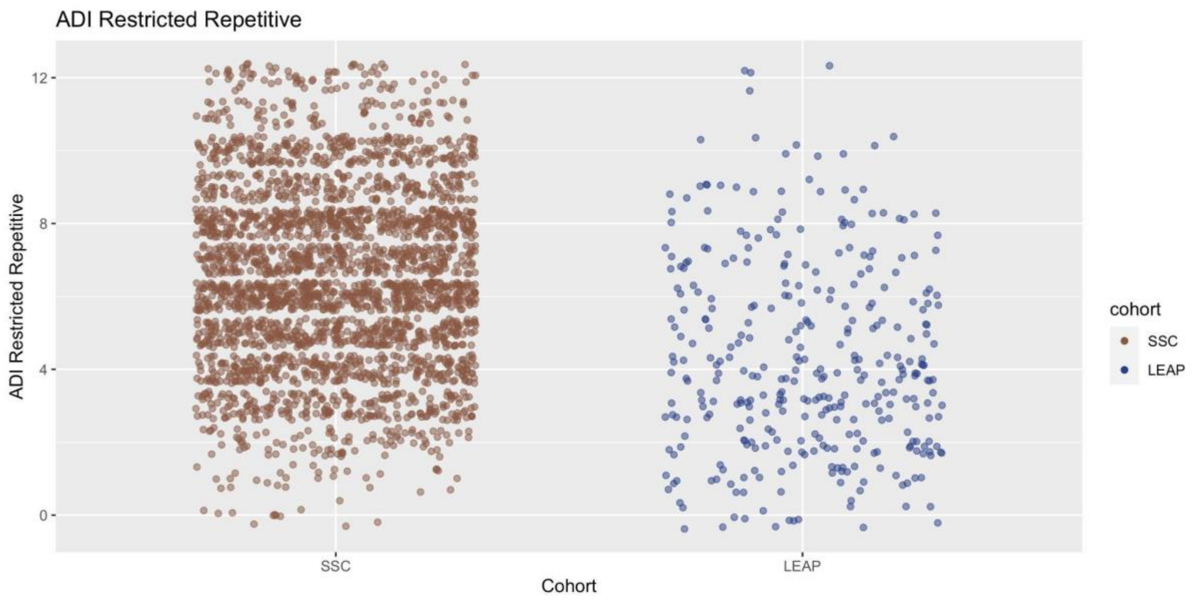


ADI-R Restricted Repetitive domain scores in all three cohorts. SSC, Simons Simplex Collection (brown); LEAP, Longitudinal European Autism Project (blue).

#### Figure S4: ADI-R Communication

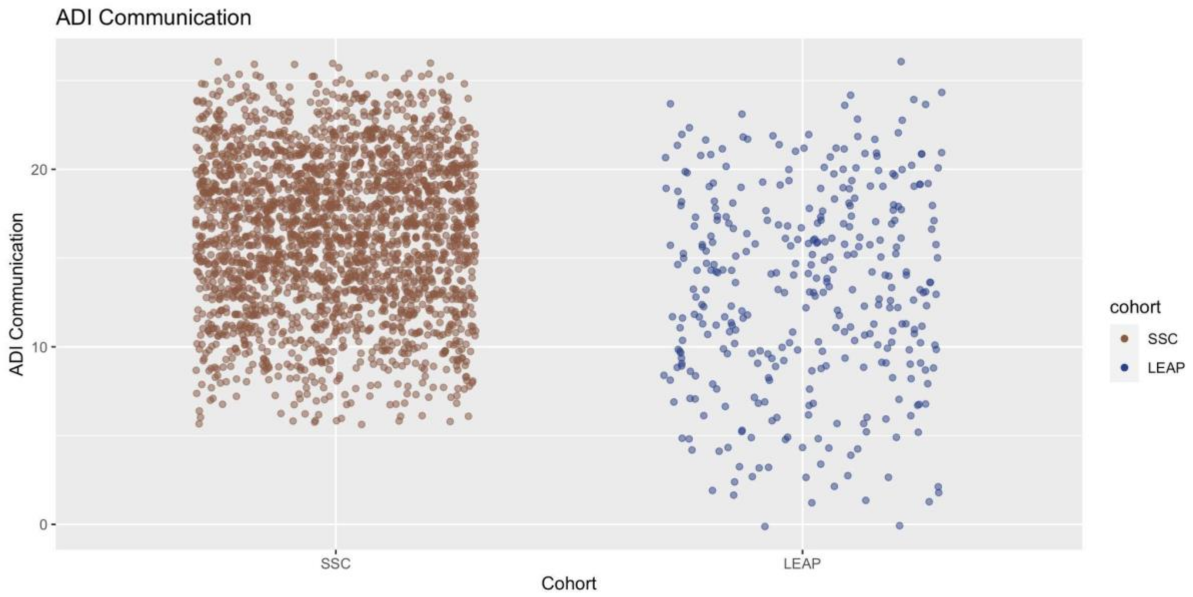


ADI-R Communication domain scores in all three cohorts. SSC, Simons Simplex Collection (brown); LEAP, Longitudinal European Autism Project (blue).

#### Figure S5: ADI-R Social

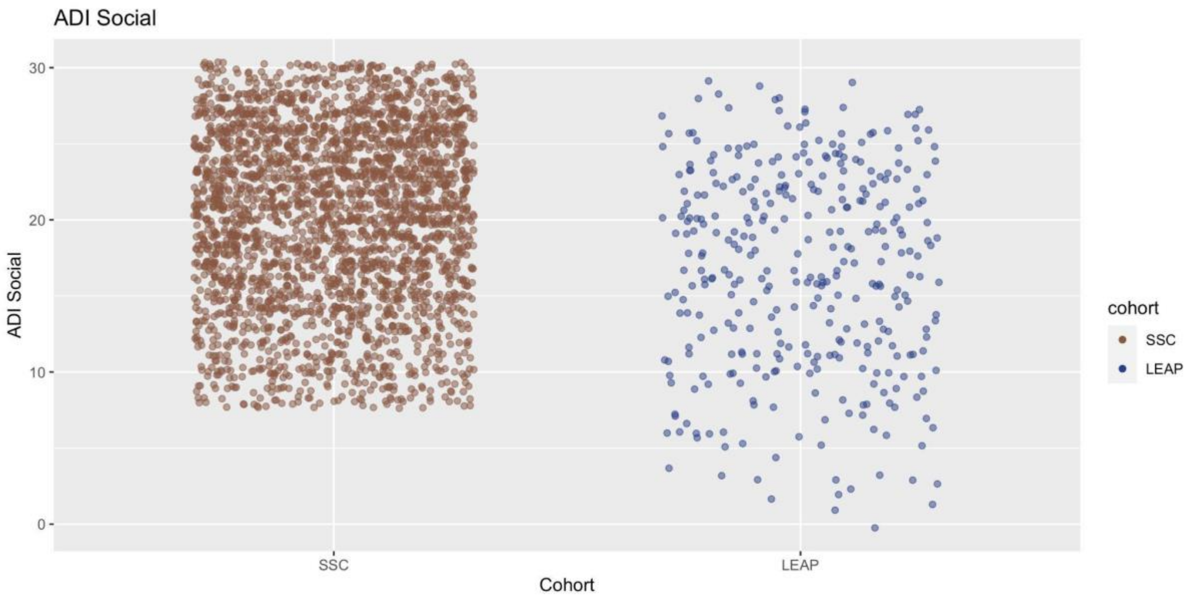


ADI-R Social domain scores in all three cohorts. SSC, Simons Simplex Collection (brown); LEAP, Longitudinal European Autism Project (blue).

Rutter, Michael, Ann Le Couteur, and Catherine Lord. 2003. “Autism Diagnostic Interview-Revised.” *Western Psychological Services*.
